# Efficacy and safety of baricitinib in patients with COVID-19 infection: Results from the randomised, double-blind, placebo-controlled, parallel-group COV-BARRIER phase 3 trial

**DOI:** 10.1101/2021.04.30.21255934

**Authors:** Vincent C. Marconi, Athimalaipet V. Ramanan, Stephanie de Bono, Cynthia E. Kartman, Venkatesh Krishnan, Ran Liao, Maria Lucia B. Piruzeli, Jason D. Goldman, Jorge Alatorre-Alexander, Rita de Cassia Pellegrini, Vicente Estrada, Mousumi Som, Anabela Cardoso, Sujatro Chakladar, Brenda Crowe, Paulo Reis, Xin Zhang, David H. Adams, E. Wesley Ely, on behalf of the COV-BARRIER Study Group

## Abstract

**Background:** The efficacy and safety of baricitinib, an oral selective Janus kinase 1/2 inhibitor, in addition to standard of care (SOC) in hospitalised adults with COVID-19 is unknown.

**Methods:** In this phase 3, global, double-blind, randomised, placebo-controlled trial, participants were enrolled from 101 centres across 12 countries in Asia, Europe, North America, and South America (ClinicalTrials.gov NCT04421027). Hospitalised adults with COVID-19 receiving SOC were randomly assigned (1:1) to once-daily baricitinib 4-mg or placebo for up to 14 days. SOC included systemic corticosteroids in 79·3% of participants (dexamethasone ∼90%). The composite primary endpoint was the proportion who progressed to high-flow oxygen, non-invasive ventilation, invasive mechanical ventilation, or death by day 28. All-cause mortality by days 28 and 60 were key secondary and exploratory endpoints, respectively. Efficacy and safety analyses included the intent-to-treat and safety populations, respectively.

**Findings:** Between June 11, 2020 and January 15, 2021, 1525 participants were randomly assigned to baricitinib 4-mg (n=764) or matched placebo (n=761). Overall, 27·8% of participants receiving baricitinib vs 30·5% receiving placebo progressed (primary endpoint, odds ratio 0·85, 95% CI 0·67-1·08; p=0·18). The 28-day all-cause mortality was 8·1% (n=62) for baricitinib and 13·1% (n=100) for placebo, corresponding to a 38·2% reduction in mortality (hazard ratio [HR] 0·57, 95% CI 0·41-0·78; nominal p=0·0018). The 60-day all-cause mortality was 10·3% (n=79) for baricitinib and 15·2% (n=116) for placebo (HR 0·62, 95% CI 0·47-0·83; p=0·0050). Frequency of serious adverse events (14·7% [n=110] vs 18·0% [n=135]), serious infections (8·5% [n=64] vs 9·8% [n=74]), and venous thromboembolic events (2·7% [n=20] vs 2·5% [n=19]) was similar between baricitinib and placebo, respectively.

**Interpretation:** While reduction of disease progression did not achieve statistical significance, treatment with baricitinib in addition to SOC (including dexamethasone) significantly reduced mortality, with a similar safety profile to SOC, in hospitalised COVID-19 participants.

**Funding:** Eli Lilly and Company.

**Research in context:** *Evidence before this study:* We searched PubMed using the terms “COVID-19”, “SARS-CoV-2”, “treatment”, “baricitinib” and “JAK inhibitor” for articles in English published up to April 31, 2020, regardless of article type. We considered previous and current clinical trials of investigational medications in COVID-19, as well as previous clinical trials of the Janus kinase (JAK)1 and JAK2 inhibitor, baricitinib, before undertaking this study. At the time the COV-BARRIER study was designed, there were no approved therapies for the treatment of COVID-19. Management of COVID-19 was supportive, and limited phase 3 randomised placebo-controlled studies had been completed. Limited phase 2 and 3 data on the antimalarial hydroxychloroquine and protease inhibitor lopinavir/ritonavir were available, and trials investigating the use of the antiviral remdesivir were ongoing. Baricitinib’s mechanism of action as a JAK1 and JAK2 inhibitor was identified as a potential intervention for the treatment of COVID-19 given its known anti-cytokine properties and potential for targeting host proteins for its antiviral mechanism. Additionally, early case series evaluating the efficacy and safety of baricitinib in the hospitalised patient population supported further evaluation of baricitinib as a potential treatment option for hospitalised patients with COVID-19. While COV-BARRIER was enrolling, ACTT-2, a phase 3 study evaluating baricitinib plus remdesivir was completed showing that baricitinib added to remdesivir improved time to recovery and other outcomes.

*Added value of this study:* This was the first phase 3 study to evaluate baricitinib in addition to the current standard of care (SOC) and included antivirals, anticoagulants, and corticosteroids. After the earliest publication of the RECOVERY study in June 2020, the treatment of hospitalised patients with COVID-19 changed with the adoption of dexamethasone as SOC. As a result of its design, COV-BARRIER became the first trial to evaluate the benefit/risk of baricitinib when added to the most current SOC (dexamethasone) in these patients. This was a randomised, double-blind, placebo-controlled trial conducted globally in regions with high COVID-19 hospitalisation rates. The reduction in the composite primary endpoint of progression to non-invasive ventilation, high flow oxygen, invasive mechanical ventilation, or death for baricitinib plus SOC (including dexamethasone) compared to placebo plus SOC did not reach statistical significance. However, in a pre-specified key secondary endpoint, treatment with baricitinib reduced 28-day all-cause mortality by 38·2% compared to placebo (HR 0·57, 95% CI 0·41-0·78; nominal p=0·0018); one additional death was prevented per 20 baricitinib-treated participants. The reduction of all-cause mortality with baricitinib was maintained by day 60 in an exploratory analysis. The frequency of serious adverse events, serious infections, and venous thromboembolic events was similar between baricitinib and placebo, respectively.

*Implications of all the available evidence:* In this phase 3 trial, baricitinib given in addition to SOC (which predominantly included dexamethasone) did not reduce a composite endpoint of disease progression, but showed a strong effect on reducing mortality by 28 days, an effect which was maintained by 60 days. In the ACTT-2 study, baricitinib further reduced time to recovery above the background use of remdesivir. Taken together, these findings suggest that baricitinib has synergistic effects to other SOC treatment modalities including remdesivir and dexamethasone. Based on all available evidence, baricitinib is a potentially effective oral treatment option to decrease mortality in hospitalised patients with COVID-19.

## INTRODUCTION

Hospitalised patients with severe acute respiratory syndrome coronavirus 2 (SARS CoV-2) often experience an intense hyperinflammatory state that may lead to multiple organ dysfunction including acute respiratory distress syndrome, septic shock, and death.^1-4^ Despite recent treatment advances with remdesivir, dexamethasone, and tocilizumab, reducing mortality among hospitalised patients remains a critical unmet need.^5-8^

Baricitinib is a selective Janus kinase (JAK)1/JAK2 inhibitor^9-11^ with a known anti-inflammatory profile in patients with autoimmune diseases.^12-14^ In February 2020, baricitinib was identified by an artificial intelligence platform as a potential intervention for the treatment of coronavirus disease 19 (COVID-19) given its known anti-cytokine properties and potential for targeting host proteins for its antiviral mechanism.^15,16^ The biochemical inhibitory effects of baricitinib on human numb-associated kinases (AAK1, BIKE, and GAK) responsible for SARS-CoV-2 viral propagation were subsequently confirmed.^17^ Baricitinib also reduced multiple cytokines and biomarkers implicated in COVID-19 pathophysiology.^18^ Following the publication of these findings, several observational studies including small cohorts of COVID-19 hospitalised patients were conducted and provided first evidence of clinical improvement associated with baricitinib treatment.^19,20^

ACTT-2, a NIH-sponsored double-blind, randomised, placebo-controlled, phase 3 trial in hospitalised adults with COVID-19, found that baricitinib plus remdesivir was superior to remdesivir in reducing time to recovery (p=0·03), with 28-day mortality reported (5·1% vs 7·8%, not statistically significant) and had fewer serious adverse events (SAEs).^6^ The Food and Drug Administration issued an Emergency Use Authorization in November 2020 for use of baricitinib, in combination with remdesivir, in hospitalised COVID-19 patients requiring oxygen supplementation.^21^ Globally, there remains a critical and urgent need to evaluate new treatment options to reduce mortality in hospitalised patients with COVID-19, because a high occurrence of deaths persists despite improvements in standard of care (SOC).

COV-BARRIER was designed to evaluate the efficacy and safety of baricitinib in combination with SOC, including dexamethasone, for the treatment of hospitalised adults with COVID-19. To the best of our knowledge, this study is the first double-blind placebo-controlled trial to evaluate mortality by day 60.

## METHODS

### Study Design and participants

This multi-centre, randomised, double-blind, placebo-controlled, parallel-group, phase 3 trial included 101 centres from 12 countries in Asia, Europe, North America, and South America. Eligible participants were ≥18 years of age, hospitalised with laboratory confirmed SARS-CoV-2 infection, had evidence of pneumonia or active, symptomatic COVID-19, and had ≥1 elevated inflammatory marker (C reactive protein, D-dimer, lactate dehydrogenase, ferritin). Participants were excluded if requiring invasive mechanical ventilation (National Institute of Allergy and Infectious Disease Ordinal Scale [NIAID-OS] 7) at study entry, receiving immunosuppressants (high dose corticosteroids, biologics, T cell or B cell-targeted therapies, interferon, or JAK inhibitors), or received convalescent plasma or intravenous immunoglobulin for COVID-19. Full inclusion and exclusion criteria are provided in the Supplementary Appendix. Following disclosure of results from ACTT-2 showing participants without baseline oxygen support were not likely to progress, a protocol amendment was implemented to limit enrollment to participants requiring baseline oxygen support (OS 5/OS 6). All participants received SOC in keeping with local clinical practice for COVID-19 management, which could include corticosteroids and/or antivirals. Dexamethasone use was permitted as described in the RECOVERY trial;^7^ higher corticosteroid doses were limited unless indicated for a concurrent condition. Prophylaxis for venous thromboembolic events (VTE) per local practice was required for all participants unless there was a major contraindication such as active bleeding events or history of heparin-induced thrombosis. The trial protocol and statistical analysis plan are available from the sponsor.

COV-BARRIER was conducted in accordance with ethical principles of the Declaration of Helsinki and Good Clinical Practice guidelines. All sites received approval from the authorized institutional review board. All participants (or legally authorized representatives) provided informed consent.

### Randomisation and masking

Randomisation was facilitated by a computer-generated random sequence using an interactive web-response system to allocate participants 1:1 to baricitinib 4-mg or placebo. Participants were stratified according to the following baseline factors: disease severity, age, region, and use of corticosteroids for COVID-19. Participants, study staff, and investigators were blinded to the study assignment. An independent, external data monitoring committee (DMC) oversaw the study and evaluated unblinded interim data for efficacy, futility, and safety analyses. An independent, blinded, clinical event committee adjudicated potential VTEs and deaths.

### Procedures

Baricitinib or placebo was administered orally (or crushed for nasogastric tube delivery) and given daily, for up to 14 days or until discharge from hospital, whichever occurred first. Participants randomised to baricitinib 4-mg with baseline eGFR ≥30 to <60 mL/min/1·73 m^2^ received baricitinib 2-mg. Efficacy and safety were evaluated for all participants up to day 28, and all cause-mortality was also evaluated in a subset of participants up to day 60. Participants had a follow-up visit ∼28 days after receiving their last dose of study drug. Efficacy and safety outcomes were assessed on scheduled study visits as indicated in the protocol. Participant disposition with reasons for discontinuation which include adverse events (AEs) (containing death) are detailed in Figure 1.

**Figure 1.**
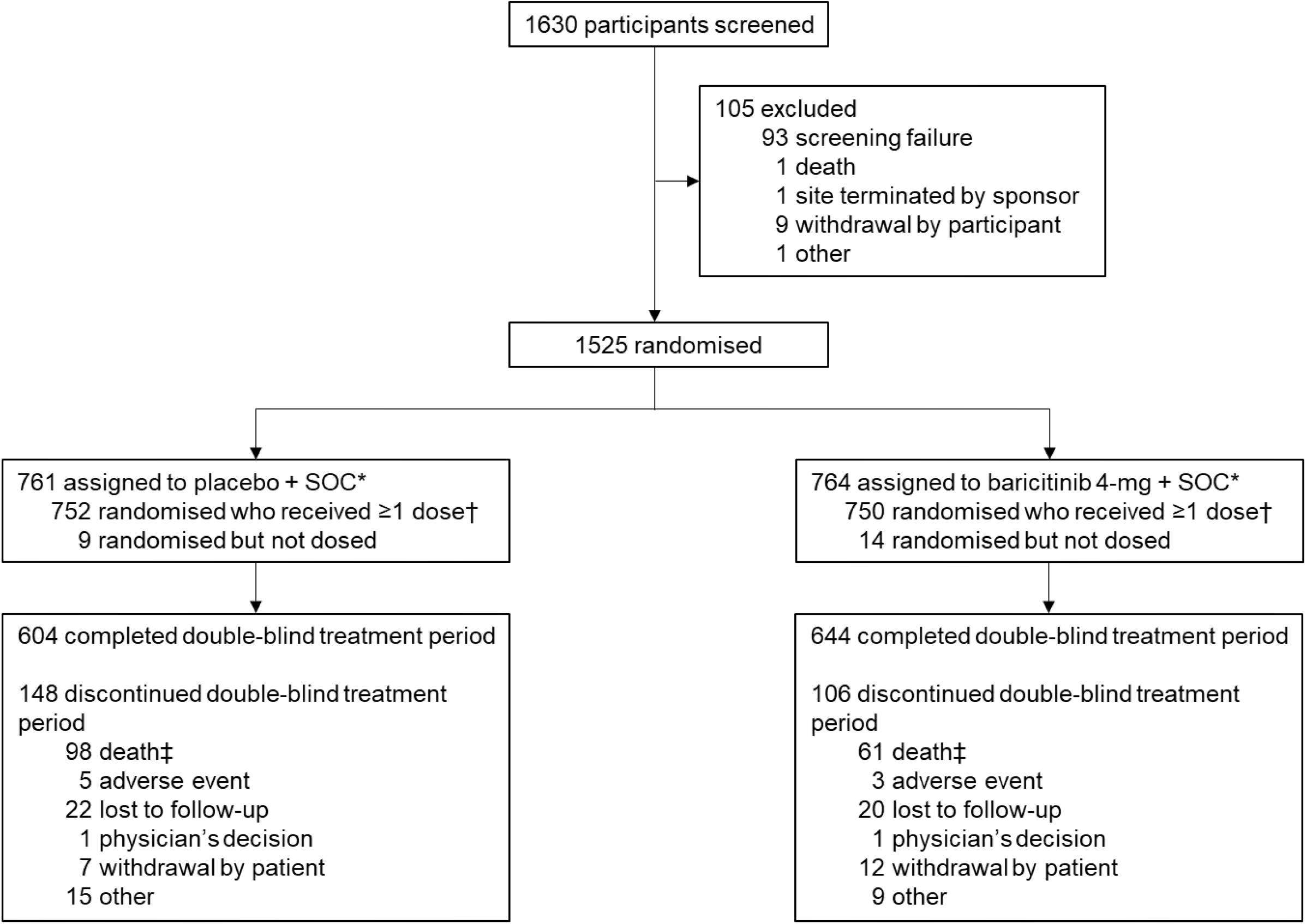
Trial profile. SOC=standard of care. *Included in the intent-to-treat population. †Includes all randomised participants who received ≥1 dose of study drug and who did not discontinue the study for the reason of ‘lost to follow-up’ at the first post-baseline visit. ‡159 deaths total reported by day 28. There were an additional three deaths that occurred after treatment period disposition but within 28 days.

### Outcomes

The composite primary endpoint was the proportion of participants who progressed to high-flow oxygen or non-invasive ventilation (OS 6), invasive mechanical ventilation or extracorporeal membrane oxygenation (OS 7), or death (OS 8) by day 28, in participants treated with baricitinib plus SOC compared to placebo plus SOC. The primary endpoint did not include outcomes subsequent to the progression during the 28-day treatment period. All deaths were assessed using the prespecified endpoint of all-cause mortality by day 28 or day 60, which was a key secondary and exploratory endpoint, respectively. The primary objective was assessed in: Population 1 – all randomised participants, and Population 2 – subpopulation of participants who, at baseline, required oxygen supplementation and were not receiving systemic corticosteroids for COVID-19.

Key secondary outcomes were adjusted for multiplicity and included the following (evaluated at days 1-28, unless otherwise specified): all-cause mortality, proportion of participants with ≥1-point improvement on NIAID-OS or discharge from hospital at days 4, 7, 10, and 14; number of ventilator-free days; time to recovery (NIAID-OS 1-3); overall improvement on the NIAID-OS evaluated at days 4, 7, 10, and 14; duration of hospitalisation; proportion of participants with a change in oxygen saturation from <94% to ≥94% from baseline to days 4, 7, 10, and 14. Pre-specified secondary outcomes and select exploratory outcomes are described in the Supplementary Appendix. Adverse events were recorded on days 1-28, coded by the Medical Dictionary for Regulatory Activities (version 23·1).

### Statistical Analyses

Power was calculated for the primary endpoint to succeed in ≥1 of the two primary populations, and the study was designed for the possibility of the sample size to be increased using an unblinded sample size re-estimation^22^ of the primary endpoint, as assessed during an interim analysis evaluated by external DMC (January 2021), where no changes were recommended. The primary and secondary endpoints were analysed according to the pre-specified analysis plan. In order to control the overall family-wise Type l error rate at a 2-sided alpha level of 0·05, a graphical testing procedure was used to test the primary and key secondary endpoints. Since the primary endpoint was not met (Table 2 and Figure S2), this manuscript reports nominal p-values for key secondary endpoints. Other analyses did not control for multiple comparisons.

Efficacy data were analysed with the intent-to-treat population, defined as all randomised participants. Logistic regression was used for dichotomous endpoints, proportional odds model was used for ordinal endpoints, analysis of variance model was used for continuous endpoints, and mixed-effects model of repeated measure was used to assess continuous endpoints over time. Log-rank test and hazard ratio (HR) from Cox proportional hazard model were used for time-to-event analyses. These statistical models were adjusted for treatment and baseline stratification factors. Safety analyses included all randomised participants who received ≥1 dose of study drug and who did not discontinue the study for the reason of ‘lost to follow-up’ at the first post-baseline visit. Adverse events were inclusive of the 28-day treatment period. Statistical tests of treatment effects were performed at a 2-sided significance level of 0·05, unless otherwise stated (i.e., graphical multiple testing strategy). Statistical analyses were performed using SAS® Version 9·4 or higher or R. Further details are described in the Supplementary Methods. This trial is registered with ClinicalTrials.gov, NCT04421027.

## RESULTS

Between June 11, 2020 and January 15, 2021, 1525 participants were randomly assigned to receive once-daily baricitinib 4-mg plus SOC (n=764) or placebo plus SOC (n=761); 83·1% (1248/1502) of participants completed the 28-day treatment period (Figure 1). Of the 16·9% (254/1502) who discontinued from the treatment period, 62·6% (159/254) of discontinuations were due to death. No randomised participants were excluded from the intent-to-treat population; however, some participants were excluded from specific analyses due to the various information requirements of the different statistical methods as outlined in Table S2.

Baseline demographics and disease characteristics were balanced among treatment groups. The mean age of the participants was 57·6 years (SD 14·1), 63·1% (n=963) were male, and enrollment was global (Table 1). Countries contributing >10% of enrollment included Brazil (22·1%, n=337), United States (21%, n=320), Mexico (18·4%, n=281), and Argentina (13·6%, n=208); participants were also enrolled in Europe, India, Japan, Korea, and Russia. Overall, 61·6% (920/1493) of participants were white, 11·7% (174/1493) were Asian, and 5·0% (75/1493) were black or African American (with 10·3% [33/320] of US participants of black or African American race). The majority (83·3% [1265/1518]) of participants had symptoms ≥7 days prior to enrollment. Clinical status at baseline was OS 4 for 12·3% (186/1518), OS 5 for 63·4% (962/1518), and OS 6 for 24·4% (370/1518) of participants. At baseline, the majority (79·3% [1204/1518]) of participants received systemic corticosteroids, of which 91·3% (1099/1204) received dexamethasone; 18·9% (287/1518) of participants received remdesivir (Table 1, Table S3). Of the participants that received remdesivir, 91·6% (263/287) also received corticosteroids. The majority of participants (99·7%, n=1520) had ≥1 pre-existing comorbid condition. In the overall population, baseline C-reactive protein value was elevated with a median value of 65·0 mg/L and was balanced between baricitinib and placebo groups (67·5 mg/L and 62·0 mg/L, respectively). Select baseline demographics and clinical characteristics by baseline systemic corticosteroid use are presented in Table S4.

**Table 1.**
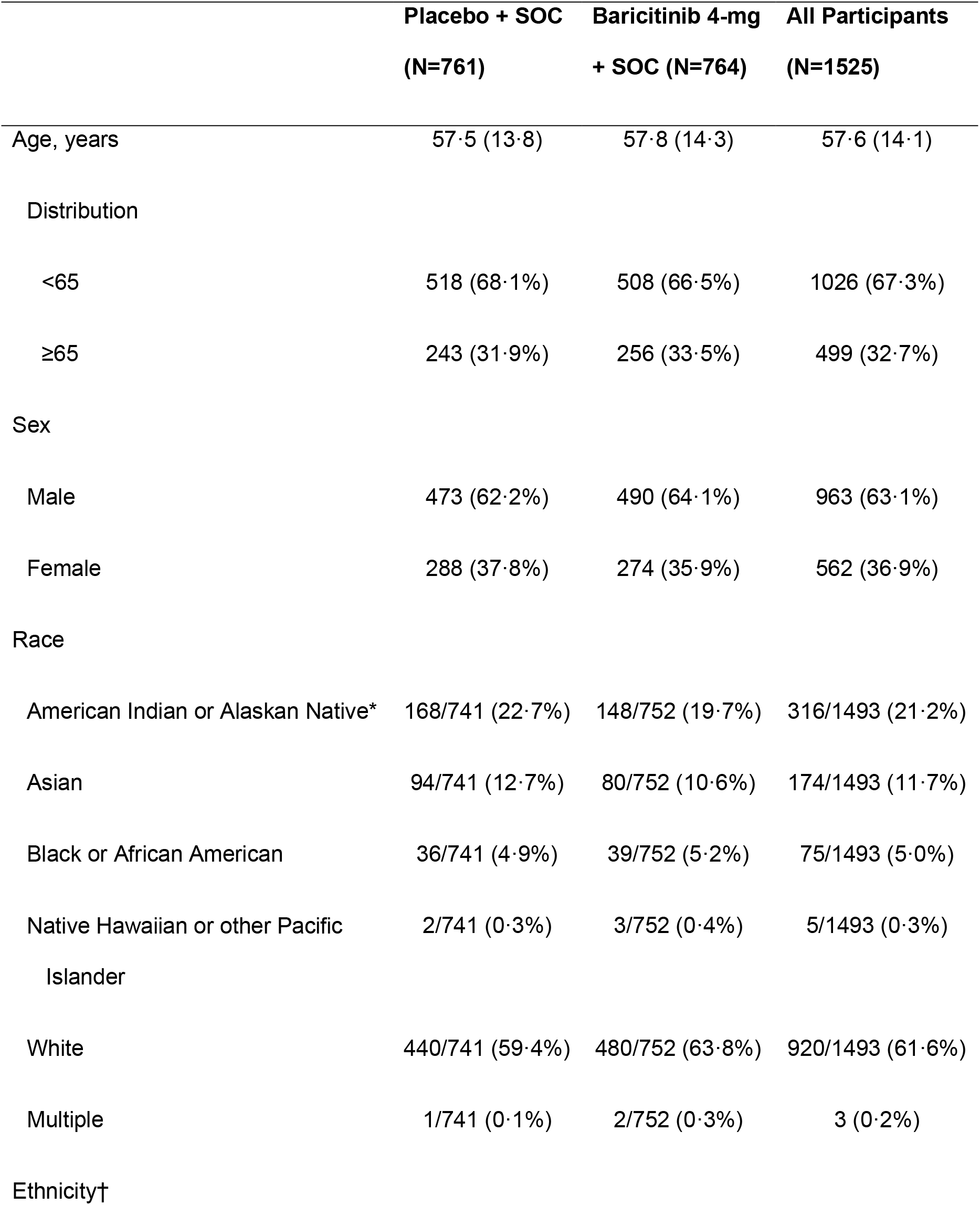
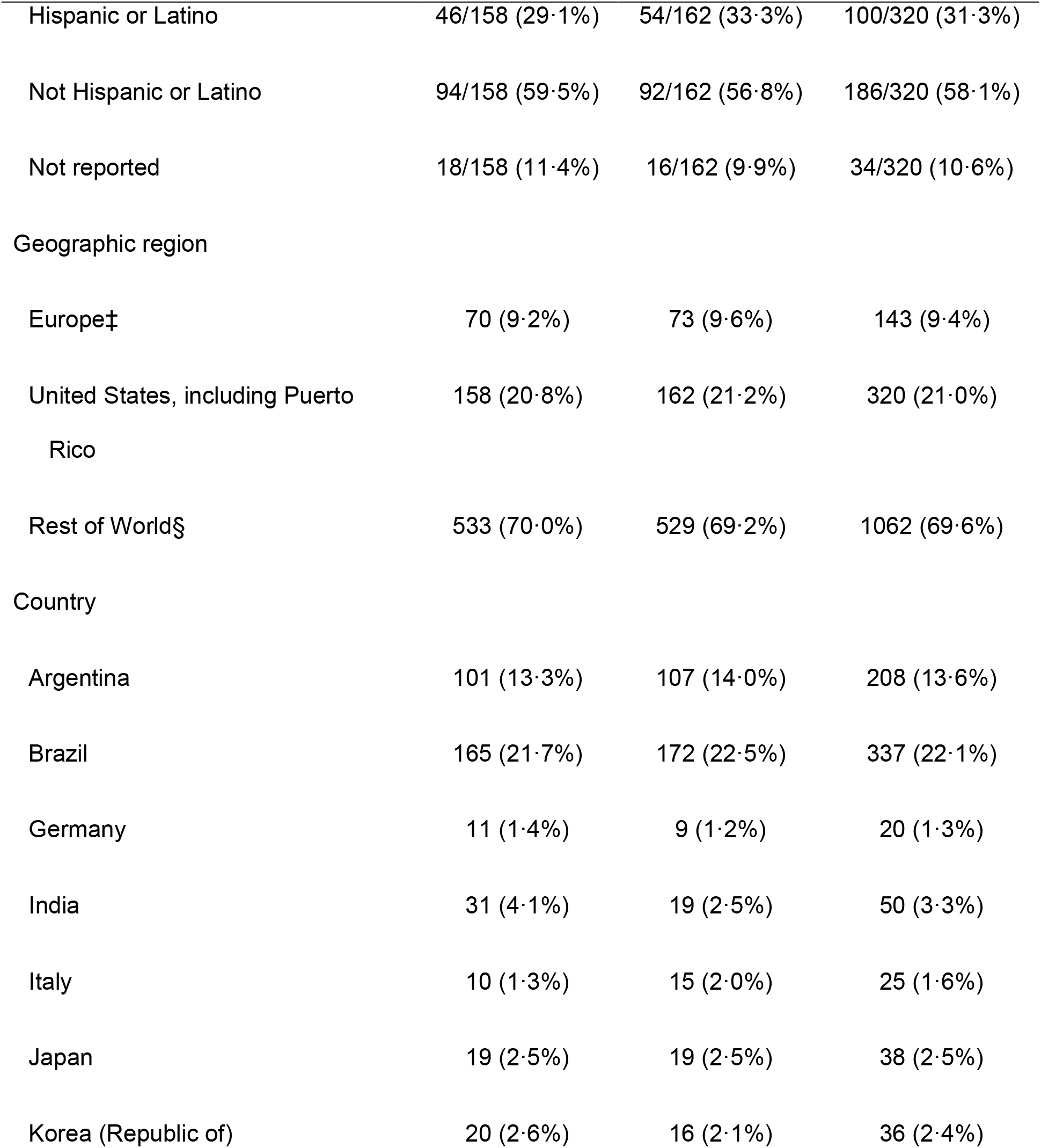
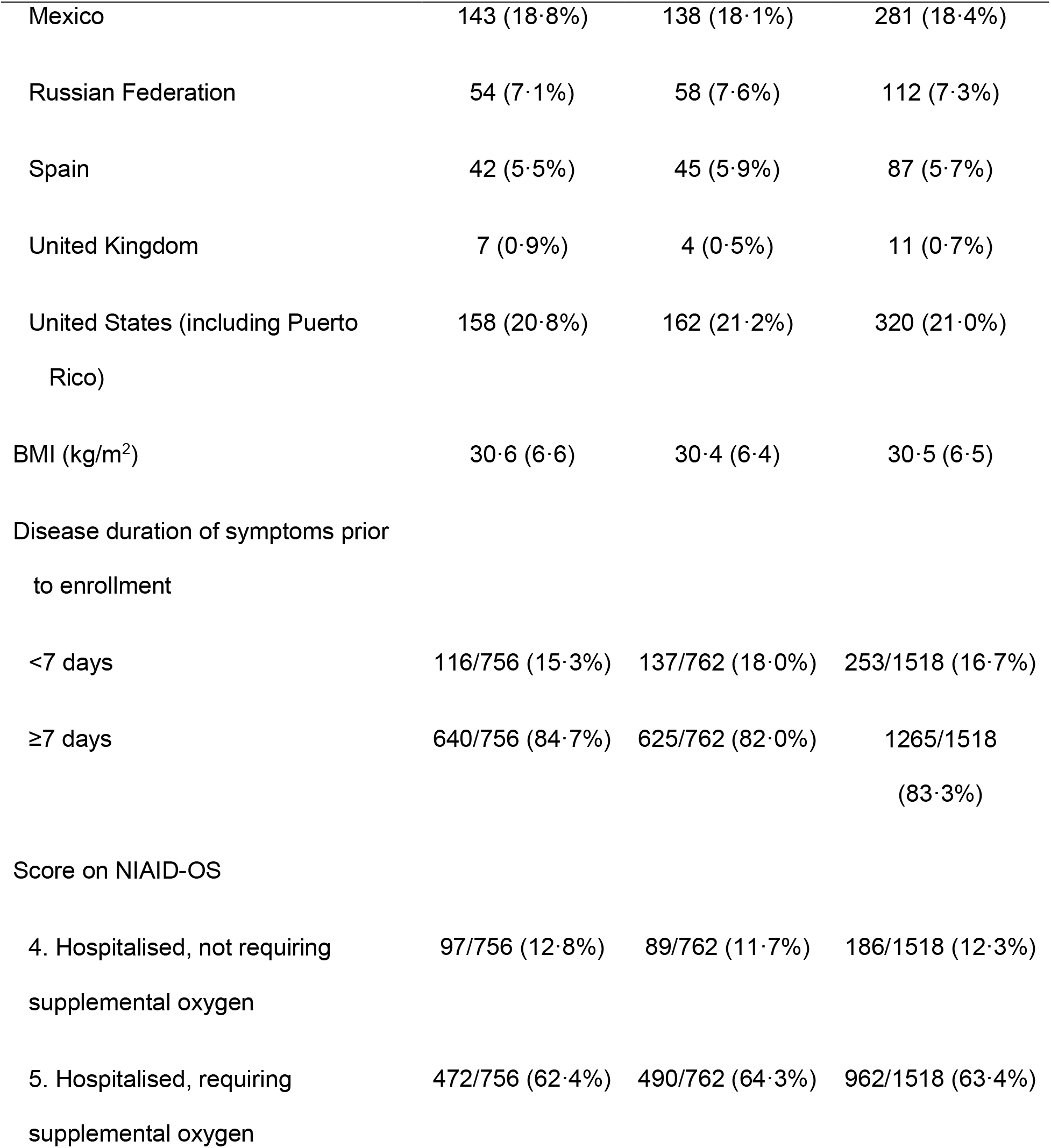
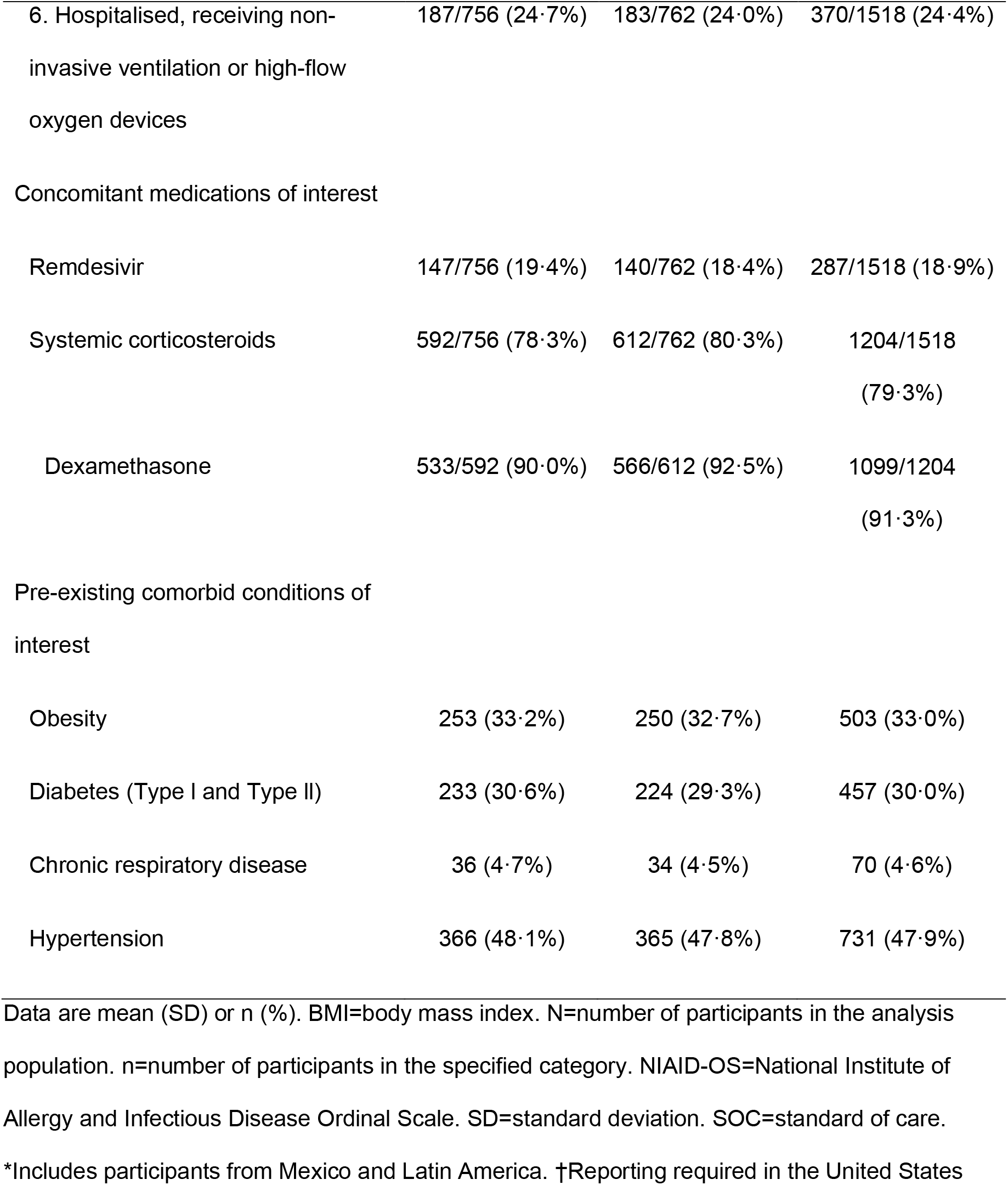
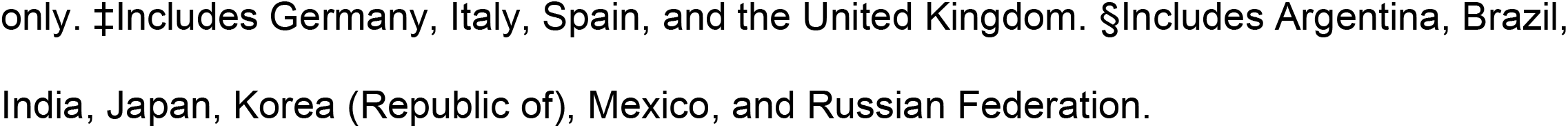
Baseline demographics and clinical characteristics.

**Table 2.**
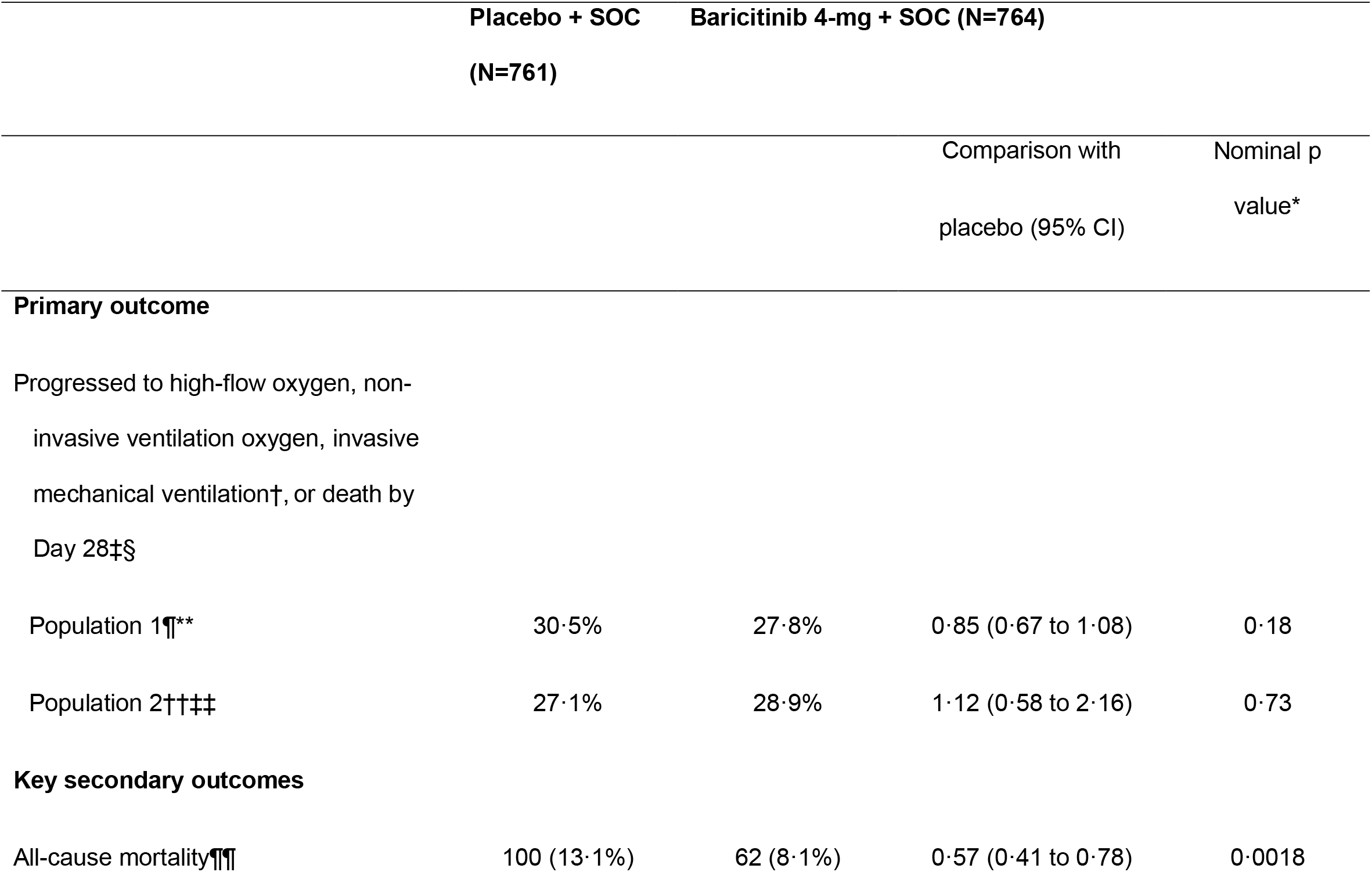
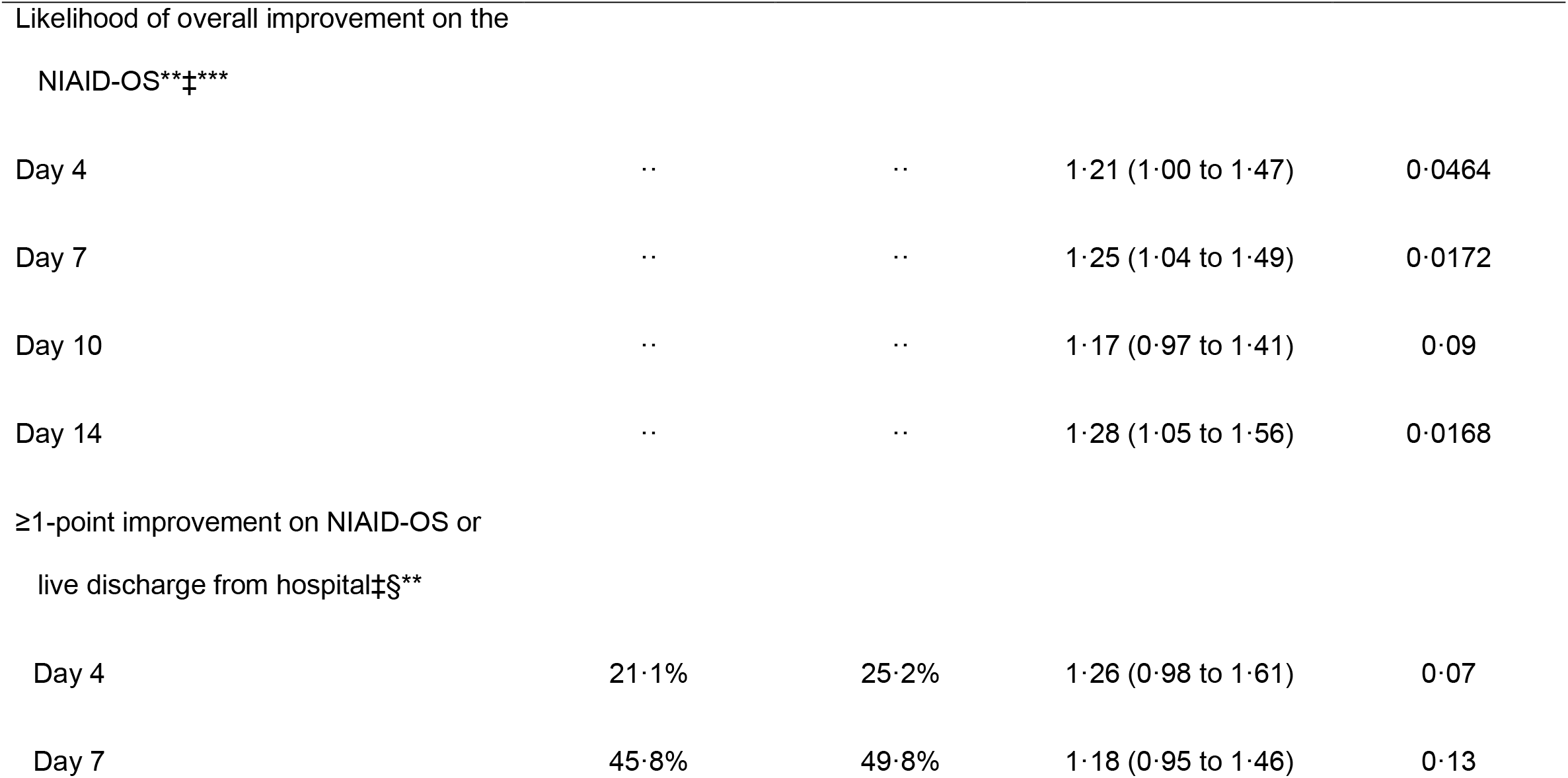
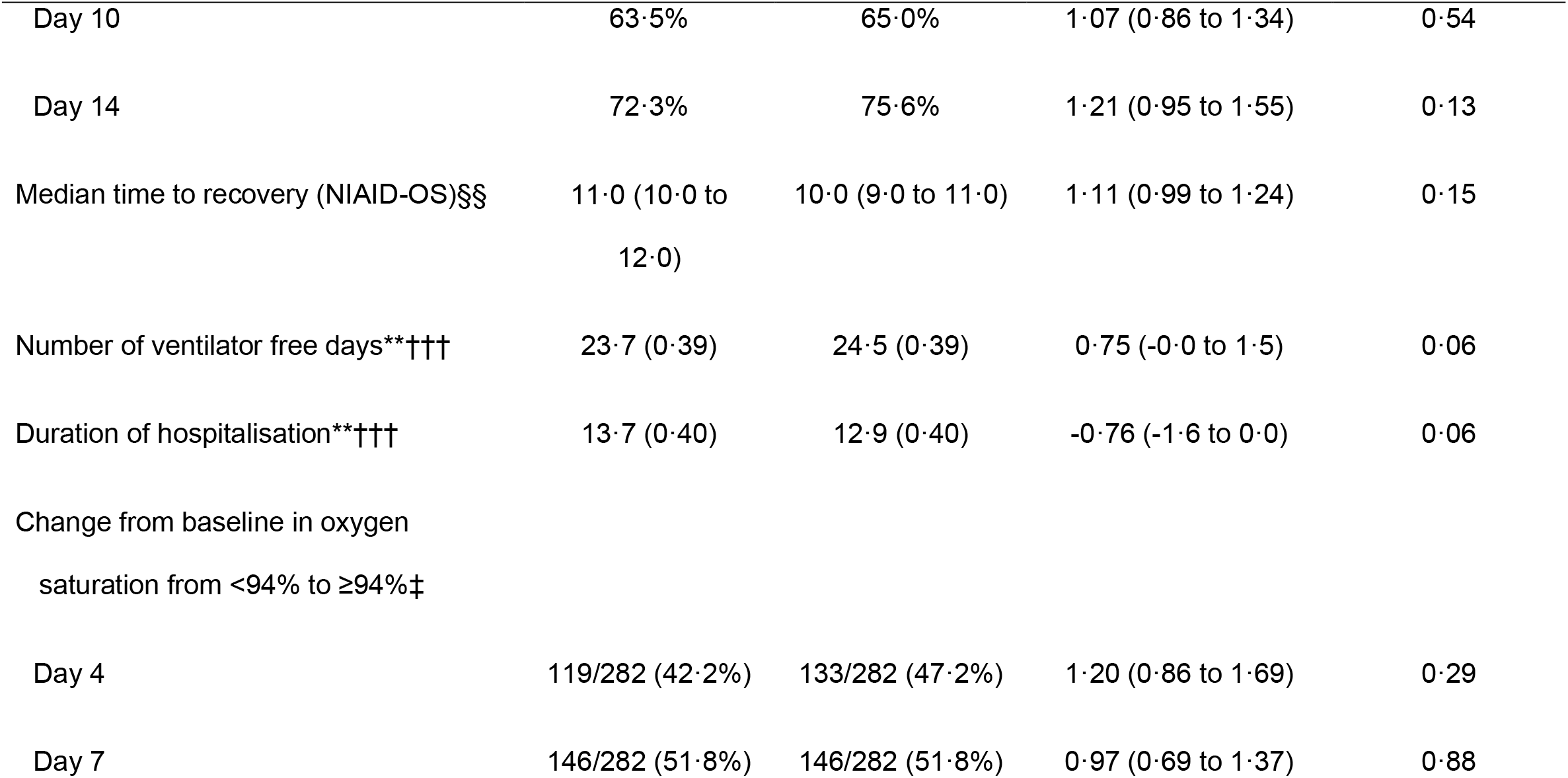
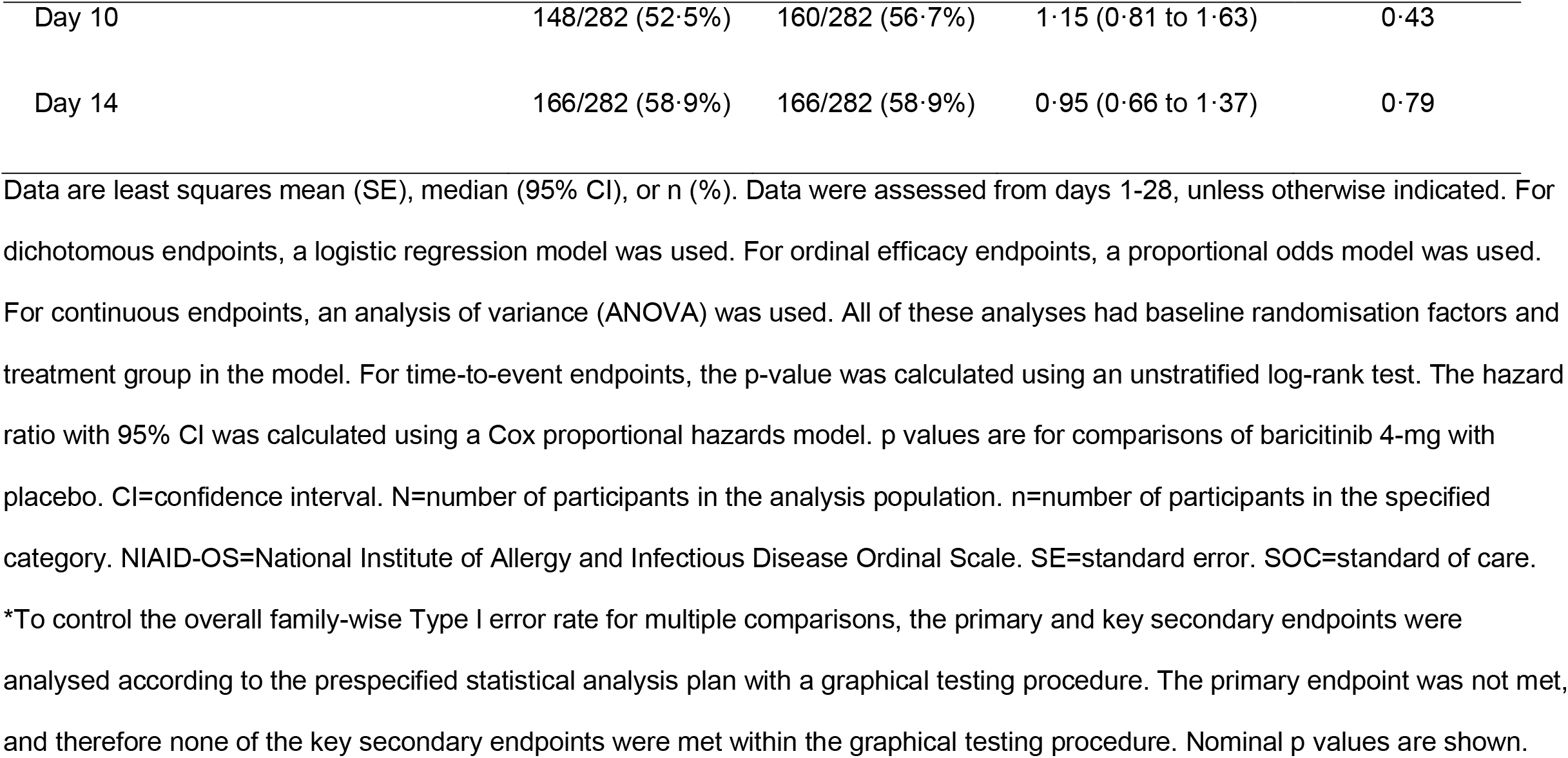
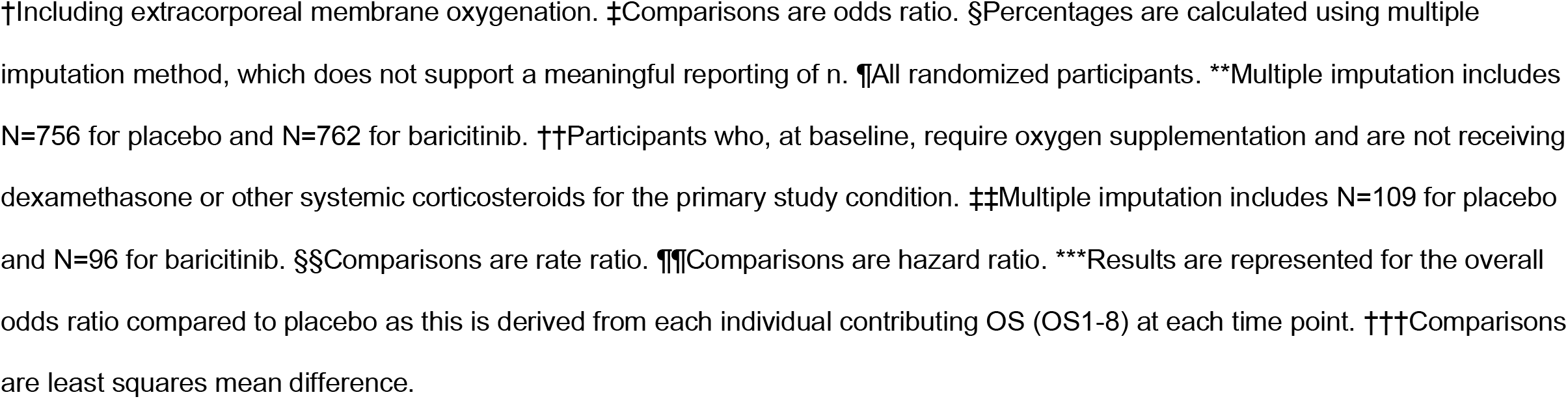
Primary and key secondary outcomes in the intent-to-treat population.

The composite primary endpoint of the proportion who progressed by day 28 revealed numerically lower progression in the group treated with baricitinib versus placebo in the analysis of Population 1 (27·8% vs 30·5%; odds ratio [OR] vs placebo 0·85, 95% confidence interval [CI] 0·67-1·08; p=0·18) without statistical significance (Table 2). Approximately 3% fewer participants progressed on baricitinib compared to placebo; this trend was observed in the overall population and across all pre-specified NIAID-OS subgroups at baseline (Table S5). In Population 2, more participants progressed with baricitinib versus placebo (28·9% vs 27·1%; OR 1·12, 95% CI 0·58-2·16; p=0·73) without statistical significance (Table 2). The primary endpoint by the pre-specified subgroup by region is reported in Table S6.

Treatment with baricitinib significantly reduced deaths, as measured by the key secondary endpoint of 28-day all cause-mortality. Mortality was 8·1% (n=62) in the baricitinib group and 13·1% (n=100) in the placebo group, corresponding to a 38·2% mortality reduction (HR 0·57, 95% CI 0·41-0·78; nominal p=0·0018); overall, one additional death was prevented per 20 baricitinib-treated participants (Table 2, Figure 2A, Figure 3). A 64·6% reduction in mortality was observed for Population 2 (5·2% [5/96] baricitinib, 14·7% [16/109] placebo; HR 0·31, 95% CI 0·11-0·88; nominal p=0·0302) (Figure 2B, Figure 3). A numerical reduction in mortality with baricitinib compared with placebo was observed for pre-specified baseline severity subgroups, including OS 4 (1·1% [1/89] vs 4·1% [4/97]; HR 0·24, 95% CI 0-2·18; nominal p=0·23) and OS 5 (5·9% [29/490] vs 8·7% [41/472]; HR 0·72, 95% CI 0·45-1·16; nominal p=0·11) (Figure 2C, Figure 3). A significant reduction in mortality was observed for baseline severity subgroup OS 6 with baricitinib compared with placebo (17·5% [32/183] vs 29·4% [55/187]; HR 0·52, 95% CI 0·33-0·80; nominal p=0·0065) (Figure 2D, Figure 3); in this subgroup, one additional death was prevented per nine baricitinib-treated participants. A significant reduction in mortality was observed with baricitinib compared with placebo for the pre-specified subgroups of participants treated at baseline with systemic corticosteroids (9·3% [57/612] vs 13·9% [82/592]; HR 0·63, 95% CI 0·45-0·89; nominal p=0·0169), without systemic corticosteroids (3·3% [5/150] vs 11·0% [18/164]; HR 0·28, 95% CI 0·10-0·77; nominal p=0·0114), or without remdesivir (8·0% [50/622] vs 13·8% [84/609]; HR 0·52, 95% CI 0·36-0·74; nominal p=0·0014); for the 18·9% (287/1518) of participants with concomitant remdesivir treatment at baseline (91·6% [263/287] also received corticosteroids), a numerical reduction in mortality was observed (Figure 2E-F, Figure S3, Figure 3). A numerical reduction in mortality was observed regardless of region for baricitinib compared with placebo (Figure 3, Table S7).

**Figure 2.**
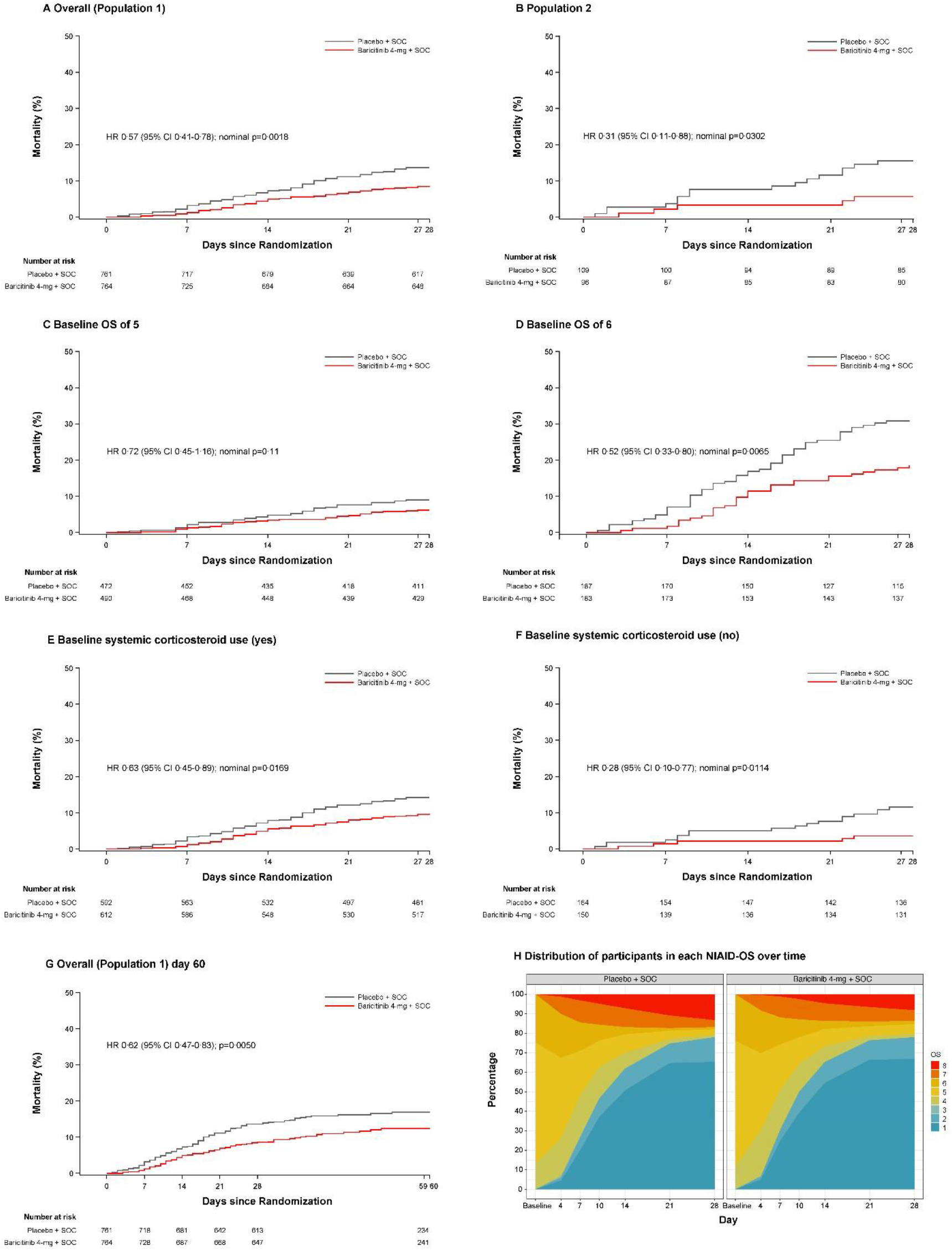
Kaplan-Meier estimates of 28-day all-cause mortality in the overall population, Population 2, by baseline NIAID-OS, and by baseline systemic corticosteroid use, 60-day all-cause mortality in the overall population, and distribution of participants in each NIAID-OS over time. For time-to-event endpoints, the p-value was calculated using an unstratified log-rank test. The HR with 95% CI was calculated using a Cox proportional hazards model. p-values are for comparisons of baricitinib 4-mg with placebo. For panels A-F, the number at risk at day 27 represent the number of participants with available data at day 28. For panel G, the number at risk at day 59 represent the number of participants with available data at day 60. Distribution of participants in each NIAID-OS over time was analysed in the intent-to-treat population with baseline OS and at least one post-baseline OS using last observation carried forward. CI=confidence interval. HR=hazard ratio. NIAID-OS=National Institute of Allergy and Infectious Disease Ordinal Scale. OS=ordinal scale. OS score of 5=hospitalised, requiring supplemental oxygen. OS score of 6=hospitalised, receiving high-flow oxygen devices or non-invasive ventilation.

**Figure 3.**
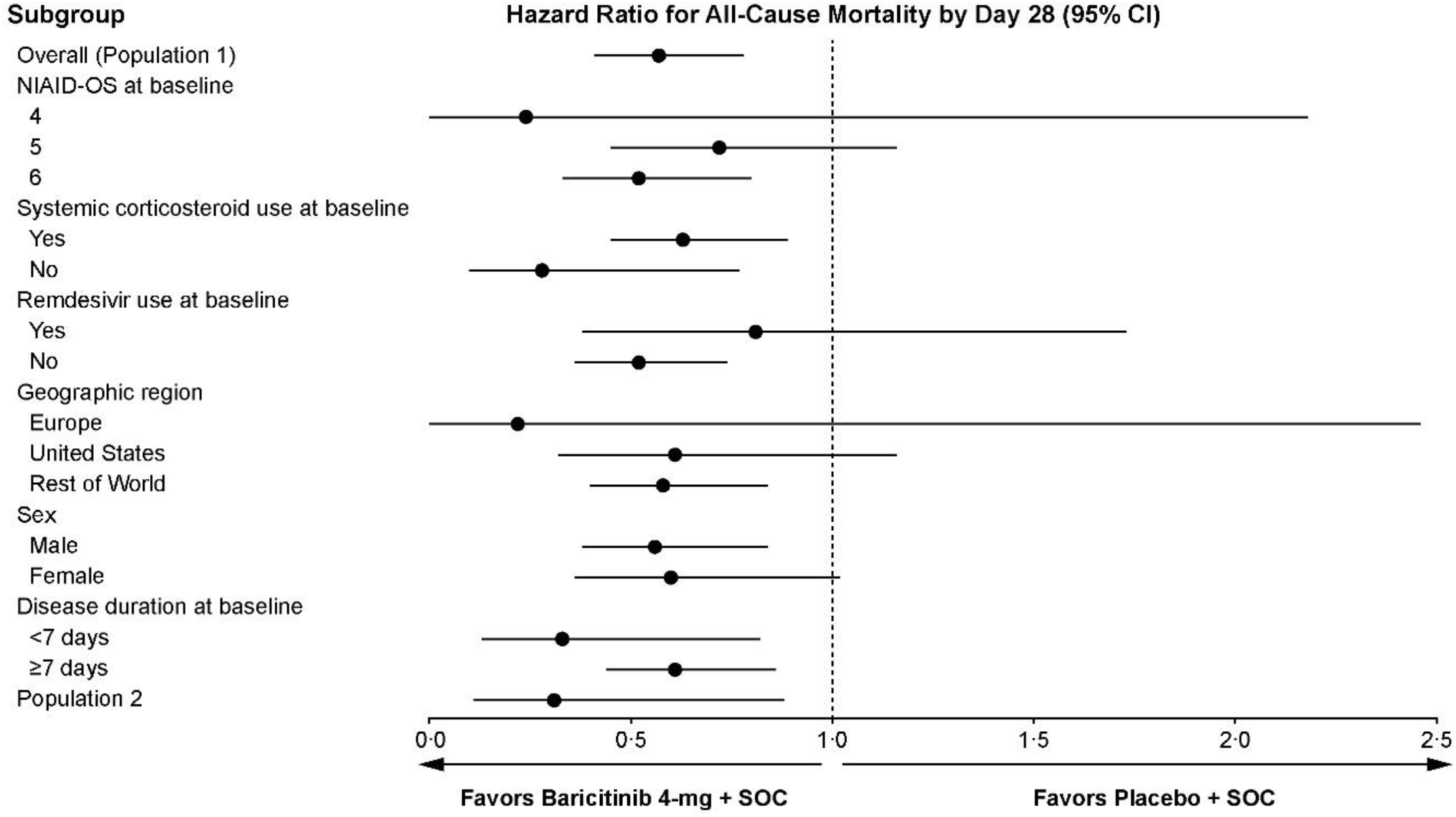
28-day all-cause mortality according to subgroup. The HR with 95% CI was calculated using a Cox proportional hazards model. CI=confidence interval. HR=hazard ratio. NIAID-OS=National Institute of Allergy and Infectious Disease Ordinal Scale.

All-cause mortality was evaluated by day 60 in a pre-specified exploratory analysis. Between day 28 and day 60, 33 additional deaths occurred in the overall population with a similar number of events in the baricitinib (n=17) and placebo (n=16) groups. The 60-day mortality remained significantly lower in the baricitinib group compared to the placebo group (10·3% [79/764] vs 15·2% [116/761]; HR 0·62, 95% CI 0·47-0·83; p=0·0050) (Figure 2G). This difference is consistent with the reduction in mortality with baricitinib by day 28. Reduced mortality was also observed for Population 2 with baricitinib treatment (5·2% [5/96] baricitinib, 17·6% [19/108] placebo; HR 0·27, 95% CI 0·10-0·75; p=0·0080) (Figure S4A). A numerical reduction in mortality with baricitinib compared with placebo was observed for the pre-specified baseline severity subgroup OS 5 (6·9% [34/490] baricitinib, 10·4% [49/472] placebo; HR 0·70, 95% CI 0·45-1·09; p=0·07) and a statistically significant reduction in mortality was observed for baseline severity subgroup OS 6 (23·0% [42/183] baricitinib, 33·7% [63/187] placebo; HR 0·58, 95% CI 0·39-0·86; p=0·0141) (Figure S4B-C). A significant reduction in mortality was observed with baricitinib compared with placebo for the pre-specified subgroups of participants treated at baseline with systemic corticosteroids (11·9% [73/612] baricitinib, 16·0% [95/593] placebo; HR 0·69, 95% CI 0·51-0·94; p=0·0440) and without systemic corticosteroids (4·0% [6/150] baricitinib, 12·9% [21/163] placebo; HR 0·30, 95% CI 0·12-0·75; p=0·0072) (Figure S4D-E).

Participant NIAID-OS status in the overall population at day 28 is shown in Figure 2H. The exploratory objective of pharmacokinetics (PK) evaluating baricitinib in participants who progressed to mechanical ventilation (intubation) and received baricitinib as a solution of crushed tablets administered via nasopharyngeal tube is shown in Figure S4. The observed PK data were most comparable to those in healthy subjects and were in the range of the PK of baricitinib 4-mg once-daily in patients with rheumatoid arthritis (Figure S5 and Supplementary Results). Other secondary endpoints are further described in the Supplementary Results, Table S8, and Figure S6.

The proportion of participants with ≥1 treatment-emergent AE (TEAE) was 44·5% [334/750] in the baricitinib group and 44·4% [334/752] in the placebo group; for SAEs, these proportions were 14·7% (n=110) and 18% (n=135), respectively (Table 3). The most common SAEs are described in Table S9. The frequency of deaths reported as due to AE (1·6% [n=12] vs 4·1% [n=31]) and discontinuation of study treatment due to AE (7·5% [n=56] vs 9·3% [n=70]) were numerically lower with baricitinib versus placebo. Serious infections were reported for 8·5% (n=64) of baricitinib-treated participants and 9·8% (n=74) of placebo-treated participants (Table 3); among participants using corticosteroids at baseline, serious infections were similar between groups (9·6% [58/605], baricitinib vs 10·7% [63/590, placebo]) (Table S10). There was a similar distribution of positively adjudicated VTEs (2·7% [n=20] vs 2·5% [n=19]) and major adverse cardiovascular events (1·1% [n=8] vs 1·2% [n=9]) with baricitinib and placebo groups, respectively. There were no reports of gastrointestinal perforations (Table 3). Safety data are described further in Tables 3, S10, and the Supplementary Results.

**Table 3.**
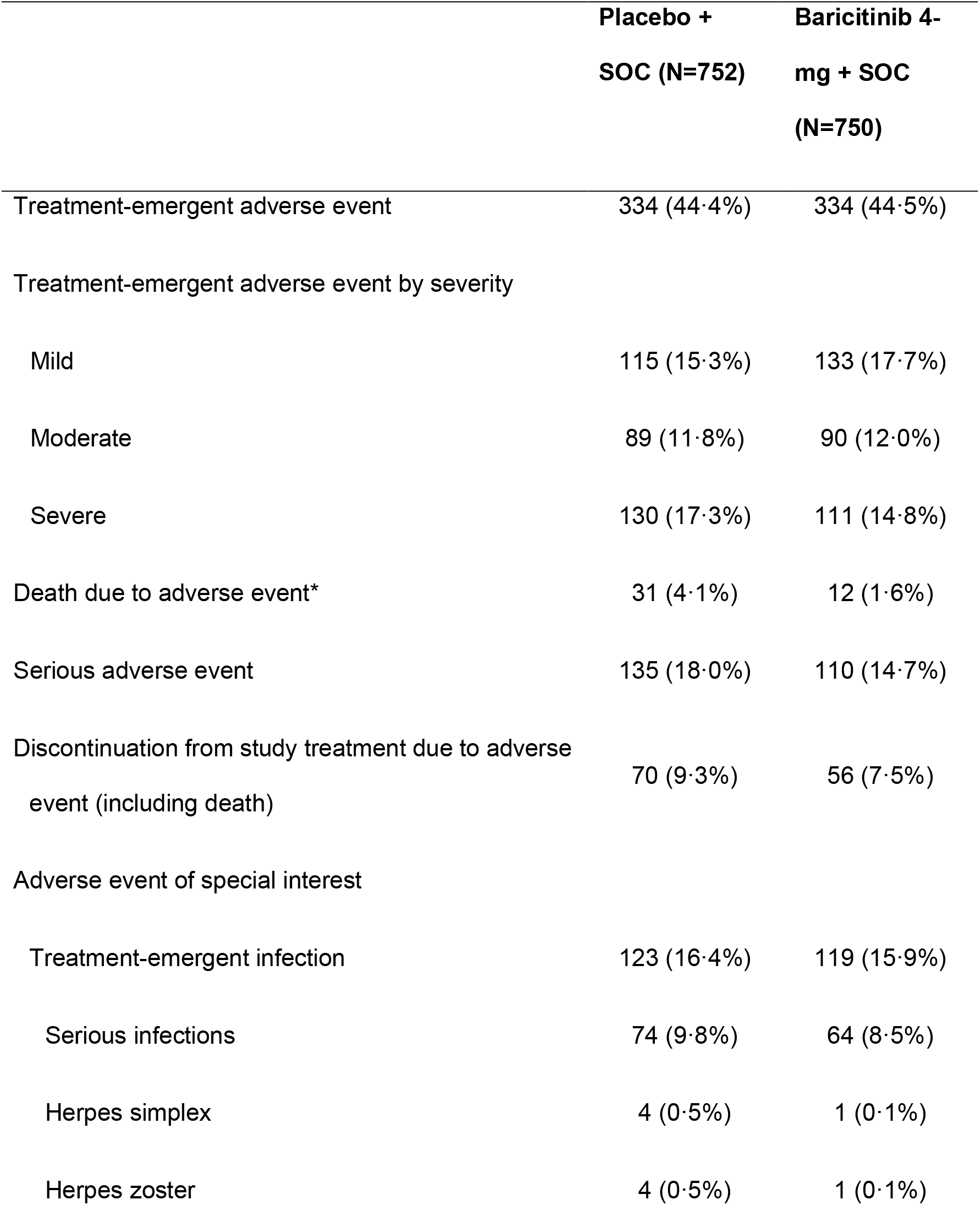
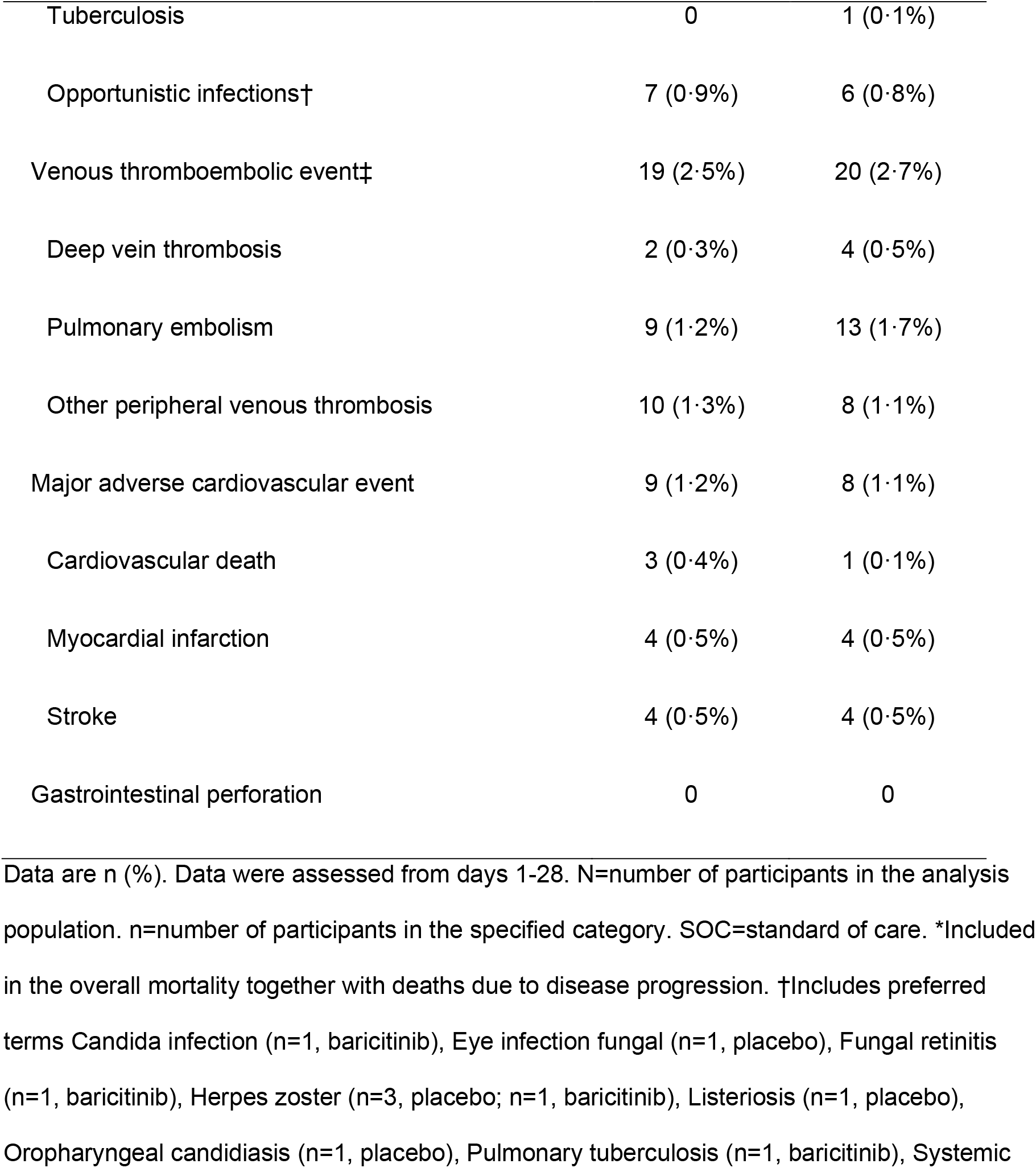
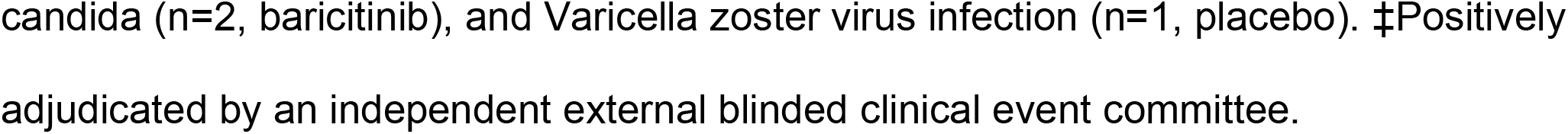
Safety overview in the safety population.

## DISCUSSION

COV-BARRIER is the first international, multi-centre, randomised, double-blind, placebo-controlled trial designed to evaluate the potential benefit and safety of baricitinib plus SOC, which included systemic corticosteroids and remdesivir, and is the first to report 60-day outcomes in a double-blind randomized controlled trial for the treatment of hospitalised adults with COVID-19. This study addresses an important knowledge gap related to the optimization of the treatment strategies for hospitalised patients with COVID-19.

In this study, the reduction of progression with baricitinib plus SOC (including dexamethasone) as defined by the composite primary endpoint did not achieve statistical significance versus placebo plus SOC. All-cause mortality is the most objective outcome of the COVID-19 disease progression in hospitalised patients and was a pre-specified key secondary endpoint of this study. Treatment with baricitinib reduced 28-day all-cause mortality by 38·2%, compared to placebo (HR 0·57, 95% CI 0·41-0·78; nominal p=0·0018); the number needed to treat (NNT) with baricitinib to prevent one additional death was 20. Mortality reduction was consistent across pre-specified subgroups of baseline disease severity and geography. In the OS 6 subgroup, the NNT with baricitinib to prevent one additional death was nine. Mortality by day 60 remained significantly lower with baricitinib plus SOC compared with placebo plus SOC, showing persistent benefit on mortality beyond the initial treatment period.

This reports the largest placebo-controlled safety data on hospitalised patients with COVID-19 treated with an immunomodulatory agent in addition to corticosteroids. The frequency of TEAEs, SAEs, infections, and VTEs were similar across the baricitinib and placebo groups and no new safety signals were detected, thus providing clinically relevant safety information for the acute care of these patients regardless of concomitant corticosteroids.

During the COV-BARRIER study, SOC changed significantly to include routine use of corticosteroids and guidelines^23,24^ were updated following the disclosure of results from the open-label RECOVERY trial in June, 2020,^25^ in which dexamethasone showed a 10·9% relative reduction in mortality compared to SOC (22·9% vs 25·7%; age-adjusted rate ratio 0·83, 95% CI 0·75-0·93; p<0·001).^7^ The evaluation of tocilizumab (anti-IL-6) in RECOVERY showed a 12·1% relative risk reduction in 28-day mortality (29% vs 33% SOC; HR 0·86, 95% CI 0·77-0·96; p= 0·007).^8^ In ACTT-2, the 28-day mortality for baricitinib plus remdesivir was 5·1% and 7·8% for remdesivir alone; the study was not powered to detect a mortality difference between the groups and the use of corticosteroids was limited.^6^ In COV-BARRIER, baricitinib plus SOC showed a 38·2% relative reduction in mortality compared to SOC including the majority of patients with use of dexamethasone at baseline (8·1% vs 13·1%; HR 0·57, 95% CI 0·41-0·78; nominal p=0·0018). To our knowledge, baricitinib showed the largest effect size on mortality for any COVID-19 treatment when compared to other randomised trials in hospitalised patients and the benefit was seen in addition to the use of background SOC (corticosteroids).^5-8^

The enrollment timeline of COV-BARRIER is also relevant considering the evolving SOC and heterogeneity of treatments across different geographies. All-cause mortality is the most relevant outcome in trials of patients hospitalised for COVID-19, and baricitinib plus SOC showed a meaningful reduction in mortality compared to placebo plus SOC, most notably for participants receiving high-flow oxygen or non-invasive ventilation.

Limitations of this study may have included the choice of disease progression, as measured by clinical status including oxygen support levels, as the primary outcome. Measuring progression based on the NIAID-OS reflects treatment decisions (such as use of certain oxygen delivery devices) and may be influenced by the heterogeneity of clinical practice across different geographies. Baricitinib showed a consistent reduction in progression versus placebo in the three components of the composite primary endpoint, however, the difference did not reach statistical significance. In contrast, mortality is an objective and definitive patient outcome that does not change across geographies. Baricitinib may prevent mortality without reaching significance on the primary endpoint because mortality as an outcome integrates the multi-organ impact of COVID-19, which include but are not limited to pulmonary effects. Baricitinib’s anti-inflammatory effects can likely have impact in organ systems that are not evaluated by the NIAID-OS.^18^ Additionally, the significant lower mortality by day 60 in the baricitinib plus SOC group versus SOC with no evaluable participants remaining on mechanical ventilation confirms that the reduction in mortality with baricitinib persists.

These results suggest that baricitinib further reduces mortality in addition to current SOC with similar safety findings and can be a treatment option to address a high unmet need in the context of the global burden of mortality observed during this COVID-19 pandemic.

## Supporting information

Marconi_VC_COV-BARRIER_Supplementary Appendix

Marconi_VC_COV-BARRIER_IRBs

Marconi_VC_COV-BARRIER_CONSORT

## Data Availability

Eli Lilly and Company provides access to all individual participant data collected during the trial, after anonymization, with the exception of pharmacokinetic or genetic data. Data are available to request 6 months after the indication studied has been approved in the US and EU and after the trial is completed, whichever is later. No expiration date of data requests is currently set once data are made available. Access is provided after a proposal has been approved by an independent review committee identified for this purpose and after receipt of a signed data sharing agreement. Data and documents, including the study protocol, statistical analysis plan, clinical study report, blank or annotated case report forms, will be provided in a secure data sharing environment. For details on submitting a request, see the instructions provided at www.vivli.org.

## Contributors

All authors contributed to the concept and design of the trial, contributed to data analysis and interpretation, critical revision of the publication, final approval to submit, and were accountable for the accuracy and integrity of the publication.

## Data sharing

Eli Lilly and Company provides access to all individual participant data collected during the trial, after anonymisation, with the exception of pharmacokinetic or genetic data. Data are available to request 6 months after the indication studied has been approved in the US and EU and after the trial is completed, whichever is later. No expiration date of data requests is currently set once data are made available. Access is provided after a proposal has been approved by an independent review committee identified for this purpose and after receipt of a signed data sharing agreement. Data and documents, including the study protocol, statistical analysis plan, clinical study report, blank or annotated case report forms, will be provided in a secure data sharing environment. For details on submitting a request, see the instructions provided at www.vivli.org.

## Acknowledgments

The authors would like to thank the participants and study investigators and staff, including the COV-BARRIER Study Group (see Supplementary Appendix) who participated in the study, and the following colleagues from Eli Lilly and Company: Douglas Schlichting, PhD (formerly with Lilly) and Maher Issa, MS, Jennifer L. Milata, PhD, Theodore Spiro, MD, and Zhongkai Wang, PhD, for scientific input, Nicole Byers, PhD for medical writing support and manuscript preparation, and Roisin McCarthy, PhD for assisting with manuscript process support.VM and JG wish to dedicate this manuscript to Francisco Marty who died this year and was an incredible champion of Infectious Diseases research and clinical care with a substantial impact on COVID-19 treatment.

