## Supplementary material for "Efficacy and safety of baricitinib in patients with COVID-19 infection: Results from the randomised, double-blind, placebo-controlled, parallel-group COV-BARRIER phase 3 trial": Marconi_VC_COV-BARRIER_Supplementary Appendix

**Table of Contents—online supplement**


| | |
|---|---|
| Page 2-5: | COV-BARRIER Study Group |
| Page 6: | Inclusion Criteria |
| Page 7-8: | Exclusion Criteria |
| Page 9-10: | Supplementary Methods |
| Page 11: | Supplementary Results |
| Page 12: | **Figure S1.** Study design |
| Page 13: | **Figure S2.** Results for graphical multiple-testing procedure |
| Page 14: | **Figure S3.** Kaplan-Meier estimates of mortality by baseline use of remdesivir |
| Page 15: | **Figure S4.** Kaplan-Meier estimates of 60-day all-cause mortality in Population 2, by baseline NIAID-OS, and by baseline systemic corticosteroid use |
| Page 16: | **Figure S5.** Pharmacokinetic profile of baricitinib 4-mg once-daily in hospitalized adults with COVID-19 |
| Page 17: | **Figure S6.** Overall improvement in the NIAID-OS evaluated at day 28 by baseline subgroup |
| Page 18: | **Table S1.** NIAID-OS |
| Page 19: | **Table S2.** Summary of populations used for different analyses |
| Page 20: | **Table S3.** Summary of systemic corticosteroids for participants with corticosteroid use at baseline |
| Page 21: | **Table S4.** Baseline demographics and clinical characteristics by baseline systemic corticosteroid use (intent-to-treat population) |
| Page 22: | **Table S5.** Proportion of participants who progressed to high-flow oxygen, non-invasive ventilation, invasive mechanical ventilation, or death (primary endpoint) per pre-specified baseline disease severity NIAID-OS subgroups by day 28 (intent-to-treat population) |
| Page 23: | **Table S6.** Proportion of participants who progressed to high-flow oxygen, non-invasive ventilation, invasive mechanical ventilation, or death by day 28 (primary endpoint) per pre-specified region subgroup by day 28 (intent-to-treat population) |
| Page 24: | **Table S7.** All-cause mortality in the intent-to-treat population by region by day 28 |
| Page 25-27: | **Table S8.** Other secondary endpoints in the intent-to-treat population |
| Page 28: | **Table S9.** Serious adverse events occurring in ≥2% of participants in either treatment group, classified by system organ class and preferred term |
| Page 29: | **Table S10.** Safety overview by baseline systemic corticosteroid use |
| Page 30: | References |

**COV-BARRIER Study Group**

**Argentina**

Javier David Altclas, Sanatorio de la Trinidad Mitre, Ciudad Autonoma de Buenos Aires, Buenos Aires

Federico Ariel, Casa Hospital San Juan de Dios, Ramos Mejía, Buenos Aires

Horacio Alberto Ariza, Clinica Central S.A., Villa Regina, Rio Negro

Anselmo Bertetti, Fundacion Sanatorio Guemes, Ciudad Autonoma de Buenos Aires, Buenos Aires

Marcelo Casas, Clinica Adventista Belgrano, Ciudad Autónoma de Buenos Aire, Buenos Aires

Valeria Cevoli Recio, Clinica Viedma, Viedma, Río Negro

Emilia Cohen, Hospital Interzonal General de Agudos "Eva Peron", San Martin, Buenos Aires

Maria Hermida, Hospital Z.G.A.D “Evita Pueblo”, Berazategui, Buenos Aires

Claudio Iastrebner, Sanatorio Sagrado Corazón, Ciudad de Buenos Aires, AR

Maria Leonor Parody, Hospital San Roque, Cordoba

Dario Scublinsky, Clinica Zabala, Ciudad de Buenos Aires, AR.

**Brazil**

Luiz Fernando Degrecci Relvas, Casa de Saude Santa Marcelina - Centro de Pesquisa Clinica, SP

Suzana Erico Tanni Minamoto, Upeclin - Unidade de Pesquisa Clínica da Faculdade de Medicina de Botucatu – UNESP, SP

Elie Fiss, Faculdade de Medicina do ABC, Santo Andre, SP

Kleber Giovanni Luz, CPCLIN, Natal, Rio Grande do Norte

Victor Augusto Hamamoto Sato, Hospital Alemão Oswaldo Cruz, SP

Maria Patelli Juliani Souza Lima, Hospital PUC-Campinas, Campinas, SP

Cesar Minelli, Hospital Carlos Fernando Malzoni Matao, Matao, SP

Priscila Paulin, CECIP - Centro de Estudos do Interior Paulista, Jaú, SP

Rita de Cassia Pellegrini, Pesquisare, Santo Andre, SP

Ana Carolina Procopio Carvalho, Hospital Felício Rocho, Belo Horizonte, Minas Gerais

Alvaro Rea Neto, CEPETI Centro de Ensino e Pesquisa em Terapia Intensiva and Centro Hospitalar de Reabilitacao Ana Carolina Moura Xavier, Curitiba, Paraná,

Cristhieni Rodrigues, Hospital Santa Paula, SP

Jose Francisco Kerr Saraiva, IPECC - Instituto de Pesquisa Clinica de Campina, Campinas, SP

Morton Scheinberg, Real e Benemerita Associação Portuguesa de Beneficiencia, SP

Anete Sevciovic Grumach, Faculdade de Medicina, Centro Universitario Saude ABC and Praxis Pesquisa Medica, Santo André, SP

Eduardo Sprinz, Hospital de Clinicas de Porto Alegre, Porto Alegre, RS

Adilson Westheimer Cavalcante, CEMEC – Centro Multidisciplinar de Estudos Clinicos EPP Ltda, São Bernardo do Campo, SP

**Germany**

Thomas Gruenewald, Klinik für Infektions- und Tropenmedizin, Klinikum Chemnitz, Chemnitz

Andreas E Kremer, Universityhospital Erlangen and Friedrich-Alexander-University Erlangen-Nürnberg, Erlangen

Stefan Schreiber, Universitätsklinikum Schleswig-Holstein, Kiel, Schleswig-Holstein

Christoph D. Spinner, Technical University of Munich, School of Medicine, University hospital rechts der Isar, Department of Internal Medicine II, Munich

**India**

Chandrasekhar Atkar, Government Medical College, Nagpur, Maharashtra

Meenakshi Bhattacharya, Government Medical College (GMC) Aurangabad, Aurangabad, Maharashtra

Akshay Budhraja, Aakash Healthcare Super Speciality Hospital, New Delhi

Kapil Zirpe, Ruby Hall Clinic and Grant Medical Foundation, Pune, Maharashtra

**Italy**

Massimo Puoti, Ospedale Niguarda Ca Granda, Milano

**Japan**

Motohiko Furuichi, Edogawa Medicare Hospital, Edogawa-ku, Tokyo

Yuji Hirai, Tokyo Medical University Hachioji Medical Center, Hachioji, Tokyo

Natsuo Tachikawa, Yokohama Municipal Citizen's Hospital, Yokohama, Kanagawa

**Korea, South**

Mi-Young Ahn, Seoul Medical Center, Seoul

Won Suk Choi, Korea University Ansan Hospital, Ansan-si, Gyeonggi-do

Jung Yeon Heo, Ajou University Hospital, Suwon, Gyeonggi-do

Eun Young Heo, Seoul National University Boramae Medical Center, Seoul, Seoul, Korea

**Mexico**

Jorge Arturo Alatorre-Alexander, Instituto Nacional de Enfermedades Respiratorias, Mexico City, México

Adrian Camacho Ortiz, Hospital Universitario "Dr. Jose Eleuterio Gonzalez", Monterrey, Nuevo León

Dora Patricia Cornejo Juarez, Instituto Nacional de Cancerologia, Mexico City, FD

Jose Guillermo Dominguez Cherit, Instituto Nacional de Ciencias Medicas y Nutricion Salvador Zubiran-Medicina Critica, FD

Michel Fernando Martínez Resendez, ITESM Campus Monterrey, Monterrey, Nuevo Leon

Gustavo Rojas Velasco, Instituto Nacional de Cardiologia Ignacio Chavez, Mexico, DF

Jesus Simon Campos, Hospital General Agustín O'Horán, Yucatan, Merida

**Russia**

Dmitry Enikeev, First Moscow State Medical University n.a. Sechenov, Moscow

Ivan Gordeev, City Clinical Hospital #15 named after O.M. Filatov, Moscow

Alexander Vishnevsky, Saint-Petersburg City Pokrovskaya Hospital, Saint-Petersburg

**Spain**

Miriam Akasbi, Hospital Universitario Infanta Leonor-Internal Medicine, Madrid

Maria Luisa Briones, Hospital Clínico Universitario de Valencia, Valencia

Carnevali Daniel, Hospital Universitario Quironsalud Madrid, Pozuelo de Alarcon, Madrid

Vicente Estrada, Hospital Clinico San Carlos, Madrid

Miguel Angel Moran Rodriguez, Hospital Txagorritxu, Vitoria, Alava

**United Kingdom**

David Hutchinson, The Royal Cornwall Hospital, Truro, Cornwall

Nidhi Sofat, Clinical Academic Group in Infection & Immunity, St George's University Hospitals NHS Foundation Trust, London

Hasan Tahir, Barnet Hospital, Barnet, Herts

**United States**

Aaliya Burza, SUNY Downstate, Brooklyn, NY

Roberto Caricchio, Temple University School of Medicine, Philadelphia, PA

Angel Comulada-Rivera, Advanced Clinical Research, LLC, Bayamon, PR

Paul Cook, East Carolina University, Greenville, NC

Todd Ellerin, South Shore Hospital, Weymouth, MA

Jason Goldman, Swedish Medical Center, Seattle, WA

Omar Gonzalez, St. Joseph's Hospital and Medical Center, Phoenix, AZ

Octavian Ioachimescu, Atlanta VA Medical Center, Decatur, GA

Manish Jain, Great Lakes Clinical Trials, Chicago, IL

Akram Khan, Oregon Health and Science University, Portland, OR

Thomas Lawrie, Sharp Memorial Hospital, San Diego, CA

Mark MacElwee, Valleywise Health, Phoenix, AZ

Farah Madhani-Lovely, Renown Regional Medical Center, Reno, NV

Vinay Malhotra, MultiCare Good Samaritan Hospital, Tacoma, WA

Patrick Milligan, Community Hospital South, Indianapolis, IN

James McKinnell, Torrance Memorial Medical Center, Torrance, CA

Priscilla Pemu, Grady Health System, Atlanta, GA

Joshua Purow, Holy Cross Hospital Inc., Fort Lauderdale, FL

Mayur Ramesh, Henry Ford Hospital, Detroit, MI

Philip Robinson, Hoag Memorial Hospital Presbyterian, Newport Beach, CA

Imad Shawa, Franciscan St. Francis Health, Indianapolis, IN

Mousumi Som, Oklahoma State University Medicine, Internal Medicine - Houston Center, Tulsa, OK

Roger Stienecker MD, Parkview Regional Medical Center, Fort Wayne, IN

Jose Suarez, Westchester General Hospital, Miami, FL

Brian Tiffany, Dignity Health Mercy Gilbert Medical Center, Gilbert, AZ

**Inclusion Criteria**

Participants are eligible to be included in the study only if they meet all of the following criteria during the screening period, unless otherwise specified below:

*Informed consent*

[1] Patient (or legally authorized representative) who gives informed consent, which includes compliance with the requirements and restrictions listed in the informed consent form (ICF) and in the protocol.

*Participant characteristics*

[2] Are male or female patients from 18 years of age (inclusive), at the time of enrollment.

Note: Contraceptive use by women should be consistent with local regulations regarding the methods of contraception for those participating in clinical studies. There are no contraceptive requirements for men.

*COVID-19 pulmonary infection-related inclusion criteria*

[3] Hospitalized with coronavirus (SARS-CoV-2) infection, confirmed by polymerase chain reaction (PCR) test or other commercial or public health assay in any specimen, as documented by either of the following:

- PCR positive in sample collected <72 hours prior to randomization; OR
- PCR positive in sample collected ≥72 hours prior to randomization (but no more than 14 days prior to randomization), documented inability to obtain a repeat sample (for example, due to lack of testing supplies, limited testing capacity, results taking >24 hours, etc.) AND progressive disease suggestive of ongoing SARS-CoV-2 infection

[4] Require supplemental oxygen at the time of study entry and at randomization.

Note: Prior to Protocol Amendment D (approved 20Oct2020) the criteria was: Have evidence of pneumonia (SpO2 <94 or PaO2/FiO2 [or SpO2/FiO2] ratio <300 mmHg or chest imaging findings consistent with pneumonia), OR have evidence of active COVID infection (with clinical symptoms including any of the following: fever, vomiting, diarrhea, dry cough, tachypnea defined as respiratory rate >24 breaths/min).

[5] Have indicators of risk of progression: at least 1 inflammatory markers >ULN (CRP, D-dimer, LDH, ferritin) with at least 1 instance of elevation >ULN within 2 days before study entry.

**Exclusion Criteria**

Participants will be excluded from study enrollment if they meet any of the following criteria within the screening period, unless otherwise specified:

*Prior or concomitant therapy*

[1] Are receiving cytotoxic or biologic treatments (such as TNF inhibitors, anti-interleukin-1 [IL-1], anti-IL-6 [tocilizumab or sarilumab], T-cell or B-cell targeted therapies (rituximab), interferon, or Janus kinase (JAK) inhibitors for any indication at study entry.

Note: A washout period 4 weeks (or 5 half-lives, whichever is longer) is required prior to screening, with the following exceptions:

- B-cell targeted therapies: a washout period of 24 weeks or 5 half-lives (whichever is longer)
- TNF inhibitors: a washout period of 2 weeks or 5 half-lives (whichever is longer), and
- JAK inhibitor: a washout period of 1 week or 5 half-lives (whichever is longer).

[2] Have ever received convalescent plasma or intravenous immunoglobulin [IVIg]) for COVID-19.

[3] Have received high dose corticosteroids at doses >20 mg per day (or prednisone equivalent) administered for ≥14 consecutive days in the month prior to study entry.

Note: Use of dexamethasone and/or other systemic corticosteroids that do not exceed the above specified dose and duration in the month prior to study entry is acceptable.

[4] Strong inhibitors of OAT3 (such as probenecid) that cannot be discontinued at study entry.

[5] Have received neutralizing antibodies, such as bamlanivimab, casirivimab and imdevimab for COVID-19.

*Current or historical infections*

Note: Documentation from verbal interview or available medical records is acceptable.

[6] Have diagnosis of current active tuberculosis (TB) or, if known, latent TB treated for less than 4 weeks with appropriate anti-tuberculosis therapy per local guidelines (by history only, no screening tests required).

[7] Suspected serious, active bacterial, fungal, viral, or other infection (besides COVID-19) that in the opinion of the investigator could constitute a risk when taking investigational product. Note: Immunocompromised patients, patients with a chronic medical condition, or those taking a medication that cannot be discontinued at enrollment, who, in the judgment of the investigator, are at increased risk for serious infections or other safety concerns given the study products should be excluded.

*Vaccines*

[8] Have received any live vaccine within 4 weeks before screening, or intend to receive a live vaccine during the study.

Note: Use of nonlive (inactivated) vaccinations is allowed for all participants.

*Other medical conditions or history*

[9] Require invasive mechanical ventilation, including ECMO at study entry.

[10] Current diagnosis of active malignancy that, in the opinion of the investigator, could constitute a risk when taking investigational product.

[11] Have a history of VTE (deep vein thrombosis [DVT] and/or pulmonary embolism [PE]) within 12 weeks prior to randomization or have a history of recurrent (>1) VTE (DVT/PE).

[12] Anticipated discharge from the hospital, or transfer to another hospital (or another unit), which is not a study site within 72 hours after study entry.

*Diagnostic assessments*

[13] Have neutropenia (absolute neutrophil count <1000 cells/μL) (<1.00 x $10^3$/μL or <1.00 GI/L)

[14] Have lymphopenia (absolute lymphocyte count <200 cells/μL) (<0.20 x $10^3$/μL or <0.20 GI/L)

[15] Have alanine aminotransferase (ALT) or aspartate aminotransferase (AST) >5 times ULN

[16] eGFR (Modification of Diet in Renal Disease [MDRD]) <30 mL/min/1.73 $m^2$.

Note: For each aforementioned diagnostic assessment (criteria 13, 14, 15, 16), 1 repeat testing is allowed during the screening period, and values resulting from repeat testing may be accepted for a participant's enrollment eligibility if the other eligibility criteria are met. In addition, these tests performed in the 24 hours prior to study entry will be accepted for determination of eligibility.

*Prior or concurrent clinical study experience*

[17] Have a known hypersensitivity to baricitinib or any of its excipients.

[18] Are currently enrolled in any other clinical study involving an investigation product or any other type of medical research judged not to be scientifically or medically compatible with this study.

Note: The participant should not be enrolled (started) in another clinical trial for the treatment of COVID-19 or SARS-CoV-2 through Day 28.

*Other exclusions*

[19] Are pregnant, or intend to become pregnant or breastfeed during the study.

[20] Are, in the opinion of the investigator or sponsor, unsuitable for inclusion in the study.

[21] Are using or will use extracorporeal blood purification (EBP) device to remove pro-inflammatory cytokines from the blood such as a cytokine absorption or filtering device, for example, CytoSorb®.

[22] Are, in the opinion of the investigator, unlikely to survive for at least 48 hours after screening.

**Supplementary Methods**

**Study design and participants**
All participants received background standard of care (SOC) in keeping with local clinical practice for COVID-19 management, which included corticosteroids (including dexamethasone), antibiotics, antivirals (including remdesivir), antifungals, and antimalarials. Dexamethasone use was permitted, consistent with the dose/duration utilized in the RECOVERY trial;[1] other corticosteroid use was limited unless indicated per SOC for a concurrent condition. Prophylaxis for venous thromboembolic events (VTE) was required for all participants unless there was a major contraindication.

**Randomization and masking**
Participants were stratified according to the following baseline stratification factors: disease severity (hospitalized not requiring supplemental oxygen, requiring ongoing medical care [National Institute of Allergy and Infectious Disease Ordinal Scale <NIAID-OS 4>; Table S1]; hospitalized requiring supplemental oxygen by prongs or mask [OS 5]; hospitalized requiring non-invasive ventilation or high-flow oxygen [OS 6]), age (<65 or ≥65 years), region (Europe, United Sates [US], or Rest of World), and use of dexamethasone and/or other systemic corticosteroid (yes/no) at baseline for COVID-19.

An independent, external data monitoring committee oversaw the study and evaluated unblinded interim efficacy and safety analyses used for safety monitoring, evaluation of excess mortality or futility, and planned sample size re-estimation. An independent, blinded, clinical event committee adjudicated potential VTEs and deaths. Full details of the trial design, conduct, oversight, amendments, and analyses are provided in the protocol and statistical analysis plan available from the sponsor.

**Other secondary and exploratory outcomes**
Prespecified secondary outcomes not adjusted for multiplicity, included the following (evaluated at days 1-28, unless otherwise specified): the time to recovery (NIAID-OS) by disease duration of <7 days or ≥7 days; duration of stay in the intensive care unit in days; time to clinical deterioration (one-category increase on the NIAID-OS); time to clinical improvement in one category of the NIAID-OS; time to resolution of fever, in participants with fever at baseline; overall improvement on the NIAID-OS evaluated at days 21 and 28; mean change in National Early Warning Score; time to definitive extubation; time to independence from non-invasive mechanical ventilation; time to independence from oxygen therapy in days; time to oxygen saturation of ≥94% on room air in days; number of days with supplemental oxygen use; number of days of resting respiratory rate <24 breaths per minute; (evaluated at days 4, 7, 10, and 14) proportion of participants in each severity category on the NIAID-OS; proportion of participants with ≥2-point improvement on the NIAID-OS or live discharge from hospital; and proportion of participants with ≥1-point improvement on the NIAID-OS or live discharge from hospital. Pre-specified exploratory outcomes included long-term (at least day 60) clinical outcomes and the characterization of the pharmacokinetics (PK) of baricitinib in intubated participants with COVID-19 infection.

**Statistical analysis**
Efficacy data were analyzed with the intent-to-treat population, defined as all randomized participants. For dichotomous and ordinal endpoints, logistic regression models, and proportional odds models were used, respectively, with baseline stratification factors and treatment group in the models. For continuous endpoints assessed at a single timepoint, analysis of variance models were used, with baseline stratification factors and treatment group in the models. For continuous measures over time, a restricted maximum-likelihood-based mixed-effects model of repeated measures was used, with treatment, baseline stratification factors, landmark days treatment-by-landmark-days-interaction as fixed categorical effects, and baseline score and baseline score-by-landmark-days-interaction as fixed continuous effects. The log-rank test was used to evaluate treatment effect in time-to-event endpoints, with Kaplan-Meier curves and median survival estimated for each treatment group. The hazard ratio (HR) with 95% confidence intervals (CI) was calculated using a Cox proportional hazards model adjusted for baseline stratification factors. Pre-specified subgroup analyses for the primary and selected key secondary endpoints evaluated treatment effect across the following subgroups: baseline OS (OS 4, OS 5, OS 6, and OS 5 and OS 6 combined), baseline usage of remdesivir (yes/no), baseline usage of corticosteroids (yes/no), region, duration of symptoms prior to enrolment, age, sex, dexamethasone and/or other systemic corticosteroid used at baseline for primary condition, and comorbidities (where applicable). Efforts to use all available data and minimize missing data imputation were considered. For time-to-event endpoints with a positive outcome (recovery or

improvement), the competing risk of death was handled by censoring participants who died at day 28. The primary missing data imputation method for endpoints related to the ordinal scale was multiple imputation using a Markov model where each transition to a future state is dependent only on the previous state. Last observation carried forward was also used to impute ordinal scales and other secondary endpoints not involving ordinal scales.

A graphical multiple-testing procedure for the primary and key secondary outcomes was prespecified to control for the Type I error rate at a two-sided alpha level of 0.05. The testing steps are as follows: first, the primary endpoint of proportion of participants who progressed to high-flow oxygen, non-invasive ventilation, invasive mechanical ventilation, or death by day 28 was tested for baricitinib plus SOC versus placebo plus SOC at a two-sided $\alpha$=0.0497 for Population 1 and at $\alpha$=0.0005 for Population 2. The two $\alpha$ were calculated accounting for the correlation between the two populations using the R package gMCP. No $\alpha$ was recycled to the key secondary outcomes as neither test for the two populations could be rejected.

### Supplementary Results

#### Select secondary outcomes

Baricitinib treatment showed improvements in select key secondary endpoints, with a statistically significant nominal p-value. Baricitinib plus SOC treatment resulted in a greater likelihood of an improvement of NIAID-OS at day 14 compared with placebo plus SOC (odds ratio [OR] 1.28, 95% CI 1.05-1.56; nominal p=0.0168), with consistent results observed at day 4 (OR 1.21, 95% CI 1.00-1.47; nominal p=0.0464), and day 7 (OR 1.25, 95% CI 1.04-1.49; nominal p=0.0172); significance was not achieved at day 10 (Table 2). Baricitinib plus SOC treatment improved other secondary outcomes not adjusted for multiplicity (Table S8). Baricitinib plus SOC treatment resulted in a higher proportion of participants that had overall improvement of OS compared to placebo plus SOC at day 7 (p=0.0274) and day 14 (p=0.0482). Among participants with fever at baseline, the median time to resolution of fever was reduced with baricitinib plus SOC compared with placebo plus SOC (3 vs 4 days, p=0.0243).

#### Safety

The proportion of participants with ≥1 TE infection was similar across groups (15.9% [119/750], baricitinib and 16.4% [123/752], placebo). Serious infections were reported for 8.5% [n=64] of baricitinib-treated participants and 9.8% [n=74] of placebo-treated participants. Herpes simplex and herpes zoster infections were reported for 1 participant (0.1%) for each infection type in the baricitinib-treated group and for 4 participants (0.5%) for each infection type in the placebo-treated group. Opportunistic infections were low in frequency and similarly distributed between baricitinib plus SOC (0.8% [n=6]) and placebo plus SOC (0.9% [n=7]), and a single tuberculosis case was reported in the baricitinib group (Table 3).

There was a similar frequency of positively adjudicated VTEs with baricitinib plus SOC (2.7% [n=20]) and placebo plus SOC (2.5% [n=19]). Deep vein thromboses (DVT) occurred in 4 participants (0.5%) in the baricitinib plus SOC group and 2 participants (0.3%) in the placebo plus SOC group. Pulmonary embolisms (PE) occurred in 13 participants (1.7%) in the baricitinib plus SOC group and 9 participants (1.2%) in the placebo plus SOC group. Other peripheral venous thromboses occurred in 8 participants (1.1%) and 10 participants (1.3%) treated with baricitinib plus SOC or placebo plus SOC, respectively. The frequency of major adverse cardiovascular events was similar with baricitinib plus SOC (1.1% [n=8]) and placebo plus SOC (1.2% [n=9]). There was 1 (0.1%) cardiovascular death reported in the baricitinib plus SOC group and 3 (0.4%) reported in the placebo plus SOC group. There were 4 (0.5%) myocardial infarctions and 4 (0.5%) strokes reported each for baricitinib plus SOC and placebo plus SOC (Table 3).

Among participants using steroids at baseline, treatment-emergent adverse events (TEAEs) and serious adverse events (SAEs) were reported for 42.8% (259/605) and 15.7% (95/605) of participants receiving baricitinib plus SOC and for 45.6% (269/590) and 19% (112/590) receiving placebo plus SOC, respectively. Deaths due to an adverse event were recorded for 1.8% (n=11) of participants in the baricitinib group and for 4.6% (n=27) of participants in the placebo group. Treatment-emergent infections were similarly reported between treatment groups (16.7% [n=101] and 16.9% [n=100]), and serious infections were reported for 9.6% (n=58) and 10.7% (n=63) of participants in the baricitinib-treated group and placebo-treated group, respectively (Table S10).

#### Pharmacokinetic characterization in adult participants with COVID-19

Plasma concentration data were available from 30 adults with COVID-19 who progressed to mechanical ventilation (intubation) and received baricitinib as a solution of crushed tablets administered via nasopharyngeal tube. These data were evaluated via graphical comparison to the known PK profiles previously characterized for other indications following a 4-mg once-daily dose administered as an oral tablet.[2,3] As shown in Figure S5, the observed PK data from participants with COVID-19 who were intubated and had baricitinib administered via nasopharyngeal tube were most comparable to those in healthy subjects and were in the range of the PK of baricitinib 4-mg once daily in patients with rheumatoid arthritis.

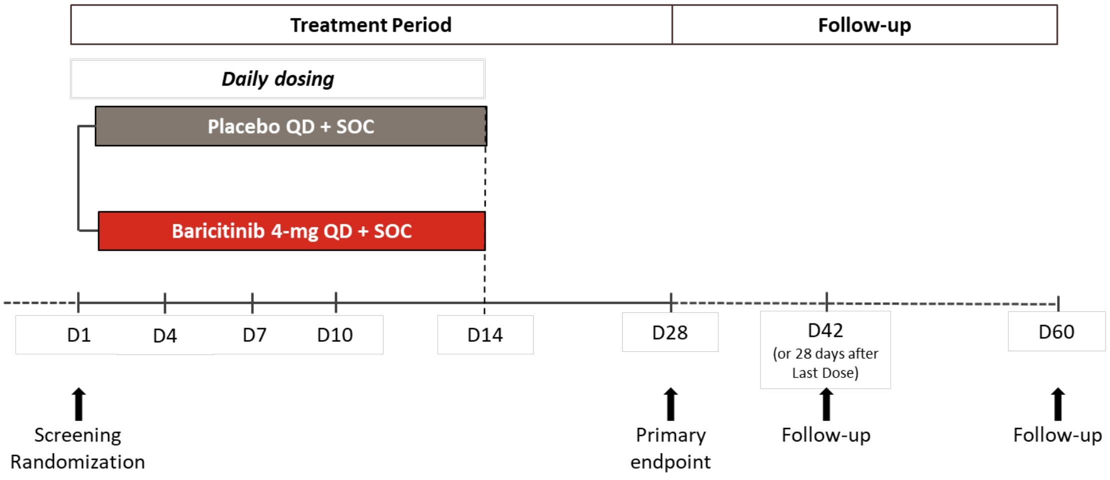


**Figure S1. Study design**
Dosing occurred from the day of randomization until day 14, or until hospital discharge or death, whichever comes first. Placebo or baricitinib 4-mg were given in addition to SOC as per local clinical practice for management of COVID-19, as defined in the protocol. D=study day. QD=once-daily. SOC=standard of care.

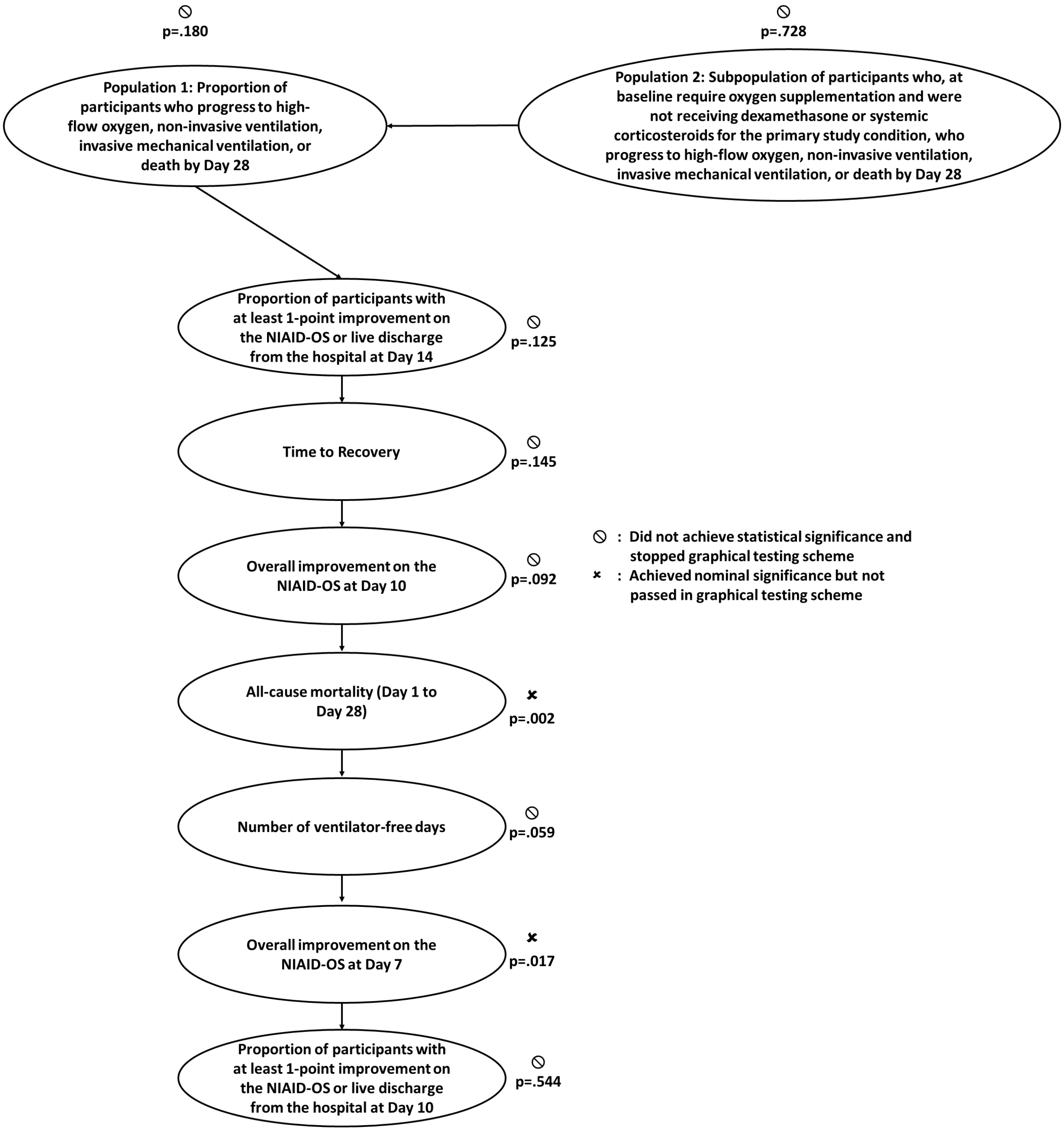


**Figure S2. Results for graphical multiple-testing procedure**
NIAID-OS=National Institute of Allergy and Infectious Diseases Ordinal Scale.

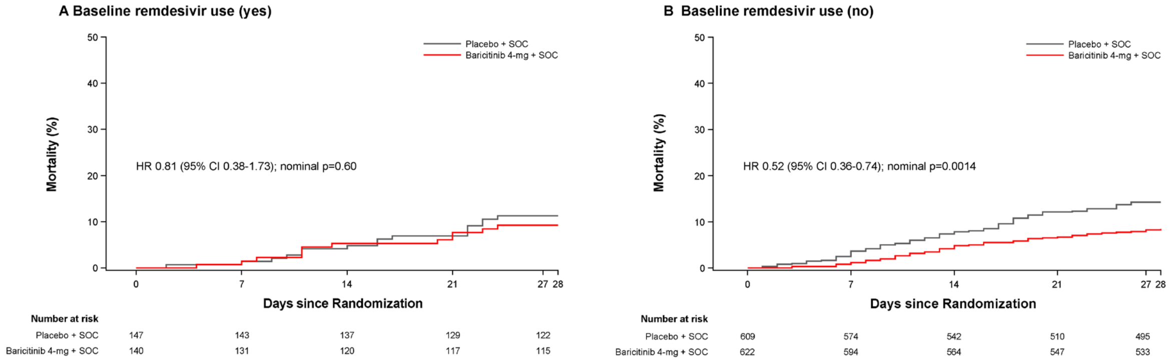


**Figure S3. Kaplan-Meier estimates of mortality by baseline use of remdesivir**
For time-to-event endpoints, the p-value was calculated using an unstratified log-rank test. The HR with 95% CI was calculated using a Cox proportional hazards model. p-values are for comparisons of baricitinib 4-mg with placebo. The number at risk at day 27 represent the number of participants with available data at day 28. CI=confidence interval. HR=hazard ratio. SOC=standard of care

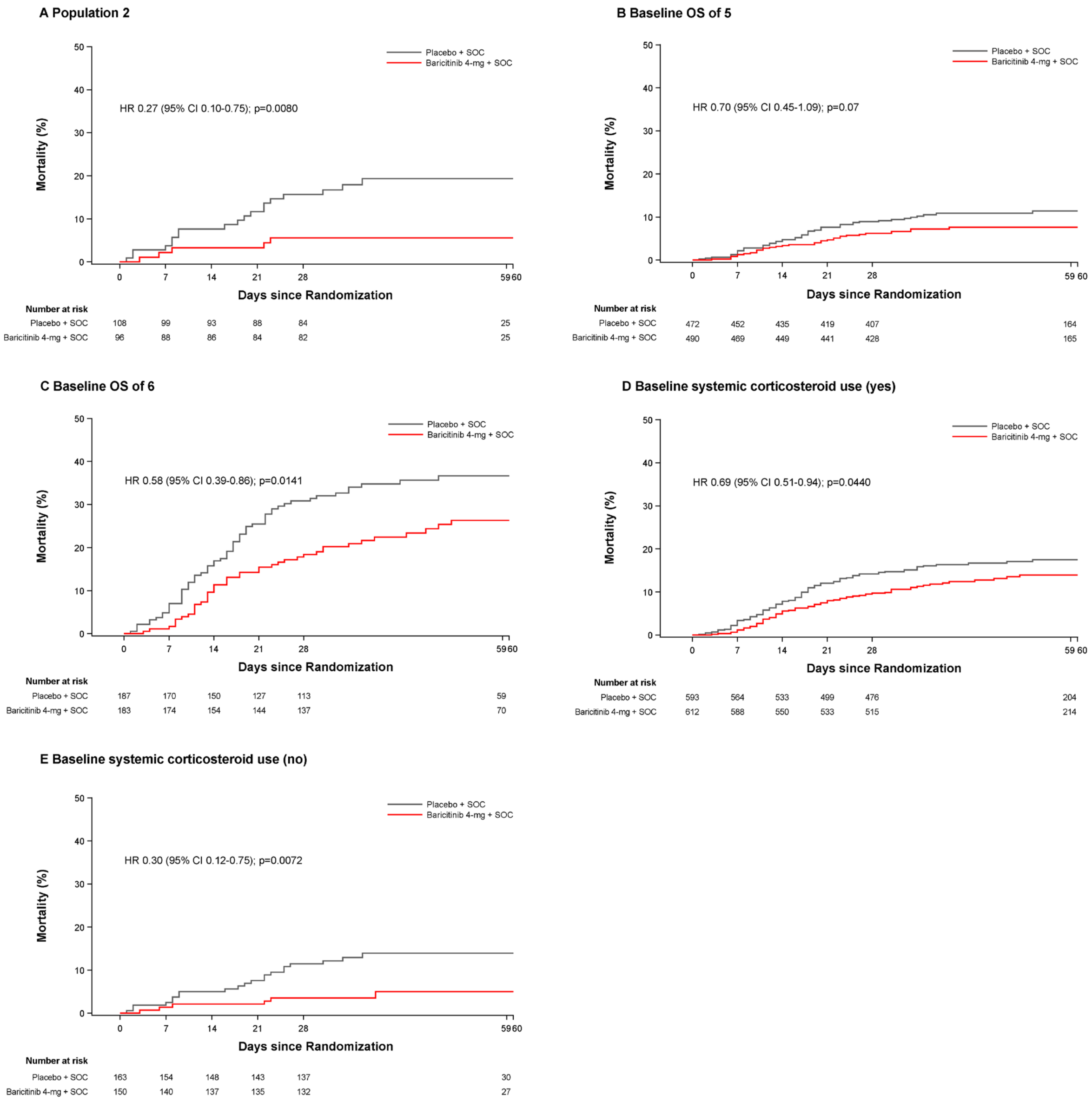


**Figure S4. Kaplan-Meier estimates of 60-day all-cause mortality in Population 2, by baseline NIAID-OS, and by baseline systemic corticosteroid use**

For time-to-event endpoints, the p-value was calculated using an unstratified log-rank test. The HR with 95% CI was calculated using a Cox proportional hazards model. p-values are for comparisons of baricitinib 4-mg with placebo. The number at risk at day 59 represent the number of participants with available data at day 60. CI=confidence interval. HR=hazard ratio. OS=ordinal scale. OS score of 5=hospitalized, requiring supplemental oxygen. OS score of 6=hospitalized, receiving high-flow oxygen devices or non-invasive ventilation. SOC=standard of care.

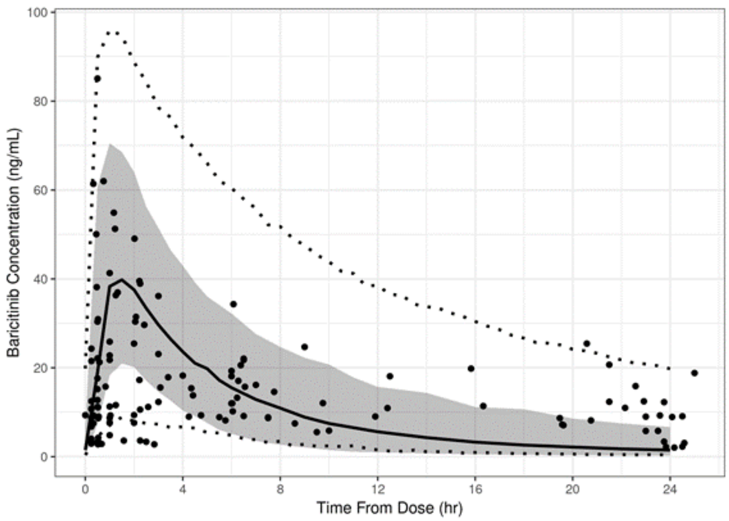


**Figure S5. Pharmacokinetic profile of baricitinib 4-mg once-daily in hospitalized adults with COVID-19**
Black symbols are observed concentration data from COV-BARRIER. Black line and grey band are model estimated median and 90% prediction interval of PK profile at 4-mg once-daily based on phase 1 clinical pharmacology studies conducted in healthy subjects. Dashed lines are model estimated 90% prediction interval of PK profiles at 4-mg once-daily baricitinib based on phase 3 studies conducted in patients with rheumatoid arthritis. PK=pharmacokinetics.

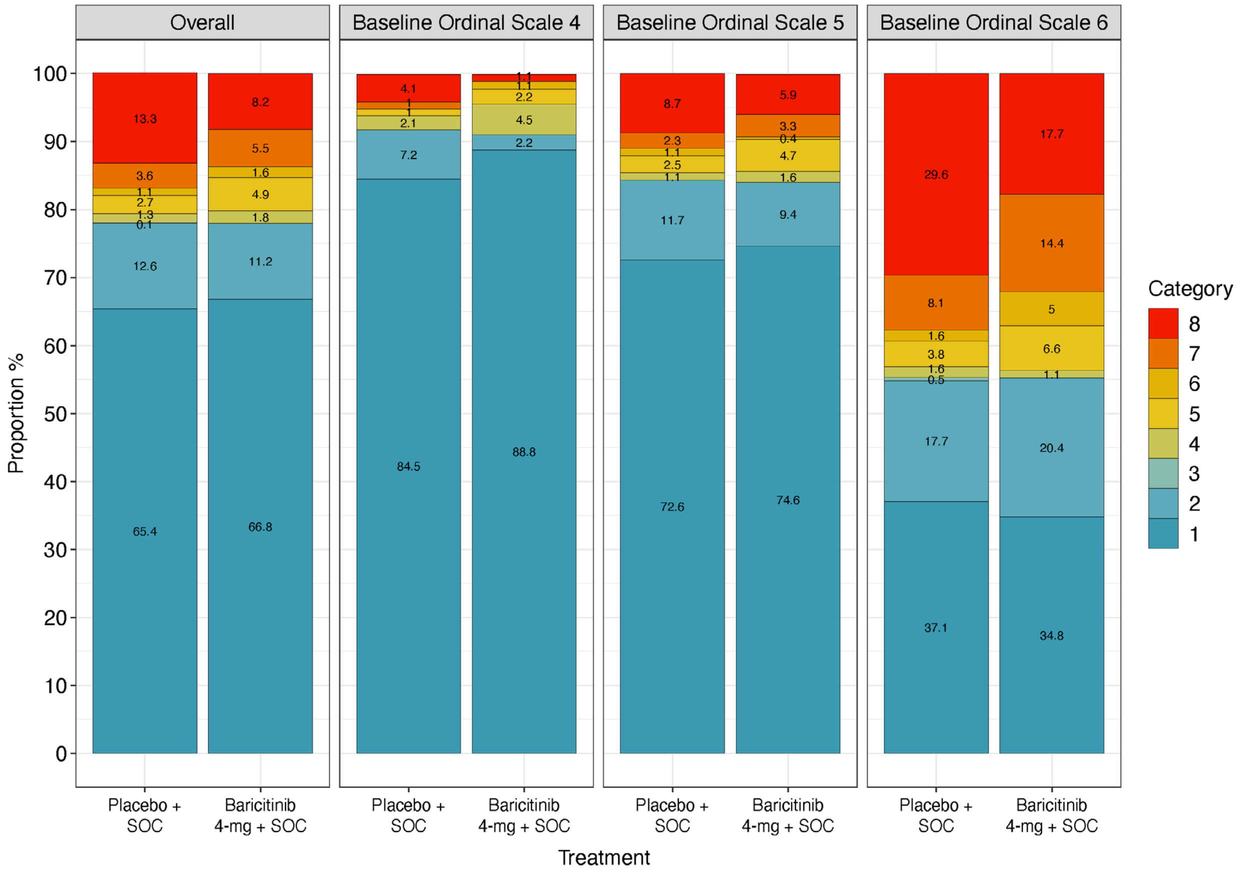


**Figure S6. Overall improvement in the NIAID-OS evaluated at day 28 by baseline subgroup.**
Intent-to-treat population with baseline OS and at least one post-baseline OS. Last observation carried forward used for analysis. NIAID-OS=National Institute of Allergy and Infectious Diseases Ordinal Scale. OS=ordinal scale. SOC=standard of care.

**Table S1. NIAID-OS**

| Score | Patient State Descriptor |
|---|---|
| OS 1 | Not hospitalized, no limitations on activities |
| OS 2 | Not hospitalized, limitation on activities and/or requiring home oxygen |
| OS 3 | Hospitalized, not requiring supplemental oxygen – no longer requires ongoing medical care |
| OS 4 | Hospitalized, not requiring supplemental oxygen – requiring ongoing medical care |
| OS 5 | Hospitalized, requiring supplemental oxygen |
| OS 6 | Hospitalized, on non-invasive ventilation or high-flow oxygen devices |
| OS 7 | Hospitalized, on invasive mechanical ventilation or extracorporeal membrane oxygenation (ECMO) |
| OS 8 | Death |

NIAID-OS=National Institute of Allergy and Infectious Disease Ordinal Scale.

**Table S2. Summary of populations used for different analyses**

| Population | Description | Total number of participants in population | Number of participants in Placebo + SOC | Number of participants in Baricitinib 4-mg + SOC | Analysis using population |
|---|---|---|---|---|---|
| ITT | All randomized participants | 1525 | 761 | 764 | All primary and key secondary Kaplan-Meier time-to-event analyses. |
| MI | All ITT participants with non-missing baseline NIAID-OS scores | 1518 | 756 | 762 | All primary and key secondary analyses involving NIAID-OS scores except for time-to-event analysis |
| LOCF | All ITT participants with non-missing baseline NIAID-OS scores and at least one non-missing post-baseline NIAID-OS score | 1512 | 754 | 758 | All secondary analyses involving NIAID-OS scores only except time-to-event analysis and analysis using MI |
| Safety | All ITT participants who receive at least 1 dose of study intervention and who were not lost to follow-up at the first postbaseline visit | 1502 | 752 | 750 | All safety analyses unless specified otherwise |

Data are N. ITT=intent-to-treat. MI=multiple imputation. LOCF=last observation carried forward. NIAID-OS=National Institute of Allergy and Infectious Disease Ordinal Scale. SOC=standard of care.

**Table S3. Summary of systemic corticosteroids for participants with corticosteroid use at baseline**

| | **Placebo + SOC (N=592)** | | | | **Baricitinib 4-mg + SOC (N=612)** | | | |
|---|---|---|---|---|---|---|---|---|
| Variable, n (%) | OS 4 (N-obs=42) | OS 5 (N-obs=384) | OS 6 (N-obs=166) | Total (N-obs=592) | OS 4 (N-obs=35) | OS 5 (N-obs=411) | OS 6 (N-obs=166) | Total (N-obs=612) |
| Beclometasone | 0 | 1 (0.3) | 2 (1.2) | 3 (0.5) | 0 | 2 (0.5) | 0 | 2 (0.3) |
| Dexamethasone | 39 (92.9) | 342 (89.1) | 152 (91.6) | 533 (90.0) | 32 (91.4) | 383 (93.2) | 151 (91.0) | 566 (92.5) |
| Hydrocortisone | 0 | 2 (0.5) | 0 | 2 (0.3) | 0 | 1 (0.2) | 0 | 1 (0.2) |
| Meprednisone | 0 | 0 | 0 | 0 | 1 (2.9) | 1 (0.2) | 0 | 2 (0.3) |
| Methylprednisolone | 2 (4.8) | 39 (10.2) | 11 (6.6) | 52 (8.8) | 2 (5.7) | 26 (6.3) | 14 (8.4) | 42 (6.9) |
| Prednisolone | 0 | 2 (0.5) | 3 (1.8) | 5 (0.8) | 1 (2.9) | 1 (0.2) | 1 (0.6) | 3 (0.5) |
| Prednisone | 1 (2.4) | 3 (0.8) | 1 (0.6) | 5 (0.8) | 0 | 1 (0.2) | 0 | 1 (0.2) |

Data are n (%). N=number of participants in the analysis population. N-obs=number of participants in the analysis. n=number of participants in the specified category. OS=ordinal scale.

**Table S4. Baseline demographics and clinical characteristics by baseline systemic corticosteroid use (intent-to-treat population)**

| | **Placebo + SOC (N=761)** | | **Baricitinib 4-mg + SOC (N=764)** | |
|---|---|---|---|---|
| Baseline systemic corticosteroid use | Yes (N-obs=592)* | No (N-obs=164)* | Yes (N-obs=612)* | No (N-obs=150)* |
| Age, years | 57.4 (13.8) | 58.2 (13.8) | 57.2 (14.0) | 59.9 (15.4) |
| Distribution, n (%) | | | | |
| <65 | 404 (68.2) | 110 (67.1) | 423 (69.1) | 84 (56.0) |
| ≥65 | 188 (31.8) | 54 (32.9) | 189 (30.9) | 66 (44.0) |
| Sex, n (%) | | | | |
| Male | 373 (63.0) | 97 (59.1) | 403 (65.8) | 87 (58.0) |
| Female | 219 (37.0) | 67 (40.9) | 209 (34.2) | 63 (42.0) |
| Score on NIAID-OS, n (%) | | | | |
| 4. Hospitalized, not requiring supplemental oxygen, requiring ongoing medical care (COVID-19-related or otherwise) | 42 (7.1) | 55 (33.5) | 35 (5.7) | 54 (36.0) |
| 5. Hospitalized, requiring supplemental oxygen | 384 (64.9) | 88 (53.7) | 411 (67.2) | 79 (52.7) |
| 6. Hospitalized, receiving non-invasive ventilation or high-flow oxygen devices | 166 (28.0) | 21 (12.8) | 166 (27.1) | 17 (11.3) |

Data are mean (SD) or n (%). N=number of participants in the analysis population. N-obs=number of participants in the analysis. n=number of participants in the specified category. NIAID-OS=National Institute of Allergy and Infectious Disease Ordinal Scale. SD=standard deviation. SOC=standard of care. *N-obs is derived from number of participants with non-missing data.

**Table S5. Proportion of participants who progressed to high-flow oxygen, non-invasive ventilation, invasive mechanical ventilation, or death (primary endpoint) per pre-specified baseline disease severity NIAID-OS subgroups by day 28 (intent-to-treat population)**

| | **Placebo + SOC (N=761)** | **Baricitinib 4-mg + SOC (N=764)** | | |
|---|---|---|---|---|
| Outcome, %* | | | Comparison with placebo OR (95% CI) | p value |
| Overall† | 30.5 | 27.8 | 0.85 (0.67 to 1.08) | 0.18 |
| NIAID-OS | | | | |
| OS 4 | 9.5 | 7.0 | 0.78 (0.27 to 2.22) | 0.64 |
| OS 5 | 28.3 | 25.6 | 0.87 (0.65 to 1.17) | 0.35 |
| OS 6 | 46.8 | 43.8 | 0.85 (0.56 to 1.30) | 0.46 |

Data are %. Data were assessed from days 1-28 using a logistic regression model with baseline randomization factors (excluding baseline disease severity for subgroups by NIAID-OS) and treatment group in the model. CI=confidence interval. N=number of participants in the analysis population. n=number of participants in the specified category. OR=odds ratio. *Percentages are calculated using multiple imputation method, which does not support a meaningful reporting of n. †Multiple imputation includes N=756 for placebo and N=762 for baricitinib.

**Table S6. Proportion of participants who progressed to high-flow oxygen, non-invasive ventilation, invasive mechanical ventilation, or death by day 28 (primary endpoint) per pre-specified region subgroup by day 28 (intent-to-treat population)**

| | **Placebo + SOC (N=761)** | **Baricitinib 4-mg + SOC (N=764)** | | |
|---|---|---|---|---|
| Outcome, n (%) | | | Comparison with placebo OR (95% CI) | p value |
| Overall* | 30.5 | 27.8 | 0.85 (0.67 to 1.08) | 0.18 |
| Region | | | | |
| Europe† | 13 (18.8) | 18 (24.7) | 1.41 (0.61 to 3.24) | 0.42 |
| United States, including Puerto Rico | 59 (37.8) | 45 (28.3) | 0.65 (0.40 to 1.06) | 0.08 |
| Rest of World‡ | 156 (29.5) | 143 (27.2) | 0.87 (0.66 to 1.16) | 0.35 |

Data are n (%). Data were assessed from days 1-28 using a logistic regression model with baseline randomization factors (excluding region for the subgroups by region) and treatment group in the model. CI=confidence interval. N=number of participants in the analysis population. n=number of participants in the specified category. OR=odds ratio. *Percentages are calculated using multiple imputation method, which does not support a meaningful reporting of n. †Includes Germany, Italy, Spain, and the United Kingdom. ‡Includes Argentina, Brazil, India, Japan, Korea (Republic of), Mexico, and Russian Federation.

**Table S7. All-cause mortality in the intent-to-treat population by region by day 28**

| Outcome, n (KM estimate %) | **Placebo + SOC (N=761)** | **Baricitinib 4-mg + SOC (N=764)** | Comparison with placebo HR (95% CI)* | p value |
|---|---|---|---|---|
| Overall | 100 (13.7) | 62 (8.6) | 0.57 (0.41 to 0.78) | 0.0018 |
| Region | | | | |
| Europe† | 4/70 (6.1) | 1/73 (1.6) | 0.22 (0 to 2.46) | 0.18 |
| United States, including Puerto Rico | 24/158 (16.6) | 16/162 (10.8) | 0.61 (0.32 to 1.16) | 0.15 |
| Rest of World‡ | 72/533 (13.8) | 45/529 (8.8) | 0.58 (0.40 to 0.84) | 0.0101 |

Data are n (KM estimate %). P-values are from a log-rank test. Hazard ratios and confidence intervals are based on Cox Proportional Hazards models with treatment and baseline stratification variables (excluding region for the subgroups by region) as explanatory variables. CI=confidence interval. KM=Kaplan-Meier. N=number of participants in the analysis population. n=number of participants in the specified category. *Favors baricitinib 4-mg + SOC if HR (95% CI) is <1. Comparisons are hazard ratio. †Includes Germany, Italy, Spain, and the United Kingdom. ‡Includes Argentina, Brazil, India, Japan, Korea (Republic of), Mexico, and Russian Federation.

**Table S8. Other secondary endpoints in the intent-to-treat population***

| Outcome | Placebo + SOC (N=761) | Baricitinib 4-mg + SOC (N=764) | Comparison with placebo (95% CI) | p value |
|---|---|---|---|---|
| Time to recovery (NIAID-OS) by baseline disease duration, days† | | | | |
| <7 days | 13.0 (10.0 to 15.0) | 11.0 (9.0 to 13.0) | 0.94 (0.70 to 1.26) | 0.22 |
| ≥7 days | 11.0 (10.0 to 11.0) | 10.0 (9.0 to 11.00) | 1.13 (1.00 to 1.28) | 0.28 |
| Duration of stay in the ICU, days‡ | 3.17 (0.31) | 3.19 (0.32) | 0.02 (-0.62 to 0.65) | 0.95 |
| Clinical deterioration (one category increase on the NIAID-OS), n (%)† | 253 (33.2) | 229 (30.0) | 0.89 (0.74 to 1.06) | 0.18 |
| Time to clinical improvement in one category of the NIAID-OS, days† | 8.0 (7.0 to 8.0) | 7.0 (7.0 to 8.0) | 1.11 (0.99 to 1.24) | 0.16 |
| Time to resolution of fever (in participants with fever at baseline), days† | 4.0 (3.0 to 4.0) | 3.0 (3.0 to 4.0) | 1.20 (1.02 to 1.42) | 0.0243 |
| Overall improvement on the NIAID-OS§¶ | | | | |
| Day 21 | .. | .. | 1.15 (0.93 to 1.43) | 0.21 |
| Day 28 | .. | .. | 1.15 (0.92 to 1.43) | 0.22 |
| Mean change from baseline in National Early Warning Score‡ | | | | |
| Day 4 | -0.59 (0.13) | -0.76 (0.13) | -0.17 (-0.42 to 0.08) | 0.19 |
| Day 7 | -0.86 (0.15) | -1.04 (0.14) | -0.17 (-0.49 to 0.14) | 0.28 |
| Day 10 | -1.33 (0.16) | -1.45 (0.16) | -0.13 (-0.49 to 0.24) | 0.50 |
| Day 14 | -1.41 (0.18) | -1.66 (0.18) | -0.25 (-0.70 to 0.19) | 0.26 |
| Definitive extubation, n (%)† | 33/136 (24.3) | 36/125 (28.8) | 1.25 (0.78 to 2.01) | 0.39 |
| Time to independence from non-invasive mechanical ventilation, days† | 11.00 (9.00 to 13.00) | 12.00 (9.00 to 14.00) | 1.09 (0.85 to 1.41) | 0.64 |
| Change in oxygen saturation from <94% to ≥94%, n (%)§** | | | | |
| Day 4 | 119/282 (42.2) | 133/282 (47.2) | 1.20 (0.86 to 1.69) | 0.29 |
| Day 7 | 146/282 (51.8) | 146/282 (51.8) | 0.97 (0.69 to 1.37) | 0.88 |
| Day 10 | 148/282 (52.5) | 160/282 (56.7) | 1.15 (0.81 to 1.63) | 0.43 |
| Day 14 | 166/282 (58.9) | 166/282 (58.9) | 0.95 (0.66 to 1.37) | 0.79 |
| Number of days with supplemental oxygen use‡ | 4.60 (0.22) | 4.37 (0.22) | -0.23 (-0.68 to 0.21) | 0.31 |
| Number of days or resting respiratory rate <24 breaths per minute‡ | 9.62 (0.30) | 9.73 (0.30) | 0.11 (-0.49 to 0.72) | 0.71 |
| Overall improvement in the NIAID-OS, n (%)§** | | | | |
| Day 4: OS at this study day | | | | |
| OS 1 | 34/754 (4.5) | 38/758 (5.0) | 1.20 (0.99 to 1.45) | 0.06 |
| OS 2 | 11/754 (1.5) | 11/758 (1.5) | | |
| OS 3 | 6/754 (0.8) | 2/758 (0.3) | | |
| OS 4 | 147/754 (19.5) | 179/758 (23.6) | | |
| OS 5 | 310/754 (41.1) | 298/758 (39.3) | | |
| OS 6 | 169/754 (22.4) | 166/758 (21.9) | | |
| OS 7 | 67/754 (8.9) | 61/758 (8.0) | | |
| OS 8 | 10/754 (1.3) | 3/758 (0.4) | | |
| Day 7: OS at this study day | | | | |
| OS 1 | 151/754 (20.0) | 187/758 (24.7) | 1.22 (1.02 to 1.47) | 0.0274 |
| OS 2 | 50/754 (6.6) | 43/758 (5.7) | | |

| | Placebo + SOC (N=761) | Baricitinib 4-mg + SOC (N=764) | | |
|---|---|---|---|---|
| Outcome | | | Comparison with placebo (95% CI) | p value |
| OS 3 | 4/754 (0.5) | 1/758 (0.1) | | |
| OS 4 | 158/754 (21.0) | 155/758 (20.4) | | |
| OS 5 | 173/754 (22.9) | 177/758 (23.4) | | |
| OS 6 | 109/754 (14.5) | 105/758 (13.9) | | |
| OS 7 | 85/754 (11.3) | 81/758 (10.7) | | |
| OS 8 | 24/754 (3.2) | 9/758 (1.2) | | |
| Day 10: OS at this study day | | | | |
| OS 1 | 280/754 (37.1) | 298/758 (39.3) | 1.14 (0.95 to 1.37) | 0.16 |
| OS 2 | 71/754 (9.4) | 81/758 (10.7) | | |
| OS 3 | 1/754 (0.1) | 1/758 (0.1) | | |
| OS 4 | 121/754 (16.0) | 108/758 (14.2) | | |
| OS 5 | 102/754 (13.5) | 103/758 (13.6) | | |
| OS 6 | 61/754 (8.1) | 69/758 (9.1) | | |
| OS 7 | 81/754 (10.7) | 79/758 (10.4) | | |
| OS 8 | 37/754 (4.9) | 19/758 (2.5) | | |
| Day 14: OS at this study day | | | | |
| OS 1 | 382/754 (50.7) | 413/758 (54.5) | 1.22 (1.00 to 1.48) | 0.0482 |
| OS 2 | 84/754 (11.1) | 81/758 (10.7) | | |
| OS 3 | 2/754 (0.3) | 1/758 (0.1) | | |
| OS 4 | 60/754 (8.0) | 61/758 (8.0) | | |
| OS 5 | 71/754 (9.4) | 67/758 (8.8) | | |
| OS 6 | 28/754 (3.7) | 31/758 (4.1) | | |
| OS 7 | 73/754 (9.7) | 68/758 (9.0) | | |
| OS 8 | 54/754 (7.2) | 36/758 (4.7) | | |
| Day 28: at this study day | | | | |
| OS 1 | 493/754 (65.4) | 506/758 (66.8) | 1.15 (0.92 to 1.43) | 0.22 |
| OS 2 | 95/754 (12.6) | 85/758 (11.2) | | |
| OS 3 | 1/754 (0.1) | 0 | | |
| OS 4 | 10/754 (1.3) | 14/758 (1.8) | | |
| OS 5 | 20/754 (2.7) | 37/758 (4.9) | | |
| OS 6 | 8/754 (1.1) | 12/758 (1.6) | | |
| OS 7 | 27/754 (3.6) | 42/758 (5.5) | | |
| OS 8 | 100/754 (13.3) | 62/758 (8.2) | | |
| ≥2-point improvement on NIAID-OS or live discharge from hospital, n (%)§** | | | | |
| Day 4 | 55/754 (7.3) | 59/758 (7.8) | 1.06 (0.72 to 1.56) | 0.78 |
| Day 7 | 215/754 (28.5) | 238/758 (31.4) | 1.14 (0.91 to 1.44) | 0.25 |
| Day 10 | 363/754 (48.1) | 385/758 (50.8) | 1.12 (0.90 to 1.38) | 0.31 |
| Day 14 | 474/754 (62.9) | 502/758 (66.2) | 1.18 (0.94 to 1.47) | 0.15 |
| Day 28 | 592/754 (78.5) | 593/758 (78.2) | 0.99 (0.76 to 1.29) | 0.93 |
| ≥1-point improvement on NIAID-OS or live discharge from hospital, n (%)§** | | | | |
| Day 4 | 158/754 (21.0) | 187/758 (24.7) | 1.23 (0.96 to 1.57) | 0.10 |
| Day 7 | 343/754 (45.5) | 369/758 (48.7) | 1.14 (0.92 to 1.40) | 0.24 |
| Day 10 | 474/754 (62.9) | 481/758 (63.5) | 1.02 (0.82 to 1.27) | 0.85 |
| Day 14 | 538/754 (71.4) | 557/758 (73.5) | 1.12 (0.89 to 1.42) | 0.33 |
| Day 28 | 604/754 (80.1) | 613/758 (80.9) | 1.06 (0.81 to 1.39) | 0.66 |

Data are median (95% CI), least squares mean (SE) or n (%). Data were assessed from days 1-28, unless otherwise indicated. For dichotomous endpoints, a logistic regression model was used. For ordinal efficacy endpoints, a proportional odds model was used. For continuous endpoints, an analysis of variance was used. All of these analyses had baseline randomization factors and treatment group in the model. For time-to-event endpoints, the p-value was calculated using an unstratified log-rank test. The hazard ratio with 95% CI was calculated using a Cox proportional hazards model. For continuous measures over time, a restricted maximum-likelihood-based mixed-effects model of repeated measures was used for comparisons with treatment, baseline randomization factors, landmark days, and treatment-by-landmark-days-interaction as fixed categorical effects, and baseline score and baseline score-by-landmark-days-interaction as fixed continuous effects. CI=confidence interval. N=number of participants in the analysis population. n=number of participants in the specified category. NIAID-OS=National Institute of Allergy and Infectious Disease Ordinal Scale. OS=ordinal scale. SE=standard error. *Prespecified objectives that were not adjusted for multiplicity. †Comparisons are hazard ratio. ‡Comparisons are least squares mean difference. §Comparisons are odds ratio; favors baricitinib 4-mg if <1. ¶Results are represented for the overall odds ratio compared to placebo as this is derived from each individual contributing OS (OS 1-8) at each time point. **Last observation carried forward used for analysis.

**Table S9. Serious adverse events occurring in ≥2% of participants in either treatment group, classified by system organ class and preferred term**

| Variable, n (%) | Placebo + SOC (N=752) | Baricitinib 4-mg + SOC (N=750) |
|---|---|---|
| Infections and infestations | 74 (9.8) | 64 (8.5) |
| COVID-19 pneumonia | 20 (2.7) | 21 (2.8) |
| Septic shock | 24 (3.2) | 13 (1.7) |
| Respiratory, thoracic and mediastinal disorders | 60 (8.0) | 43 (5.7) |
| Acute respiratory failure | 29 (3.9) | 17 (2.3) |
| Respiratory failure | 17 (2.3) | 10 (1.3) |

Data are n (%). N=number of participants in the analysis population. n=number of participants in the specified category.

**Table S10. Safety overview by baseline systemic corticosteroid use**

| | **Placebo + SOC (N=752)** | | **Baricitinib 4-mg + SOC (N=750)** | |
|---|---|---|---|---|
| Baseline systemic corticosteroid use | Yes (N-obs=590) | No (N-obs=162) | Yes (N-obs=605) | No (N-obs=145) |
| Treatment-emergent adverse event | 269 (45.6) | 65 (40.1) | 259 (42.8) | 75 (51.7) |
| Death due to adverse event | 27 (4.6) | 4 (2.5) | 11 (1.8) | 1 (0.7) |
| Serious adverse event | 112 (19.0) | 23 (14.2) | 95 (15.7) | 15 (10.3) |
| Treatment-emergent infection | 100 (16.9) | 23 (14.2) | 101 (16.7) | 18 (12.4) |
| Serious infections | 63 (10.7) | 11 (6.8) | 58 (9.6) | 6 (4.1) |

Data are n (%). N=number of participants in the analysis population. N-obs=number of participants in the analysis. n=number of participants in the specified category.
