## Supplementary material for "Efficacy and safety of baricitinib in patients with COVID-19 infection: Results from the randomised, double-blind, placebo-controlled, parallel-group COV-BARRIER phase 3 trial": Marconi_VC_COV-BARRIER_IRBs

COV-BARRIER IRBs and Approval Dates

| **Country** | **Site** | **IRB Name** | **Approval Date** |
| --- | --- | --- | --- |
| United States | 100 | Western Institutional Review Board - Connexus | 06-Jun-2020 |
| United States | 101 | Western Institutional Review Board - Connexus | 05-Jun-2020 |
| United States | 102 | South Shore Hospital IRB | 10-Jun-2020 |
| United States | 103 | Western Institutional Review Board - Connexus | 14-Jul-2020 |
| United States | 104 | Western Institutional Review Board - Connexus | 15-Jun-2020 |
| United States | 106 | Sharp Center for Research Institutional Review Board | 20-Jul-2020 |
| United States | 108 | Western Institutional Review Board - Connexus | 02-Jun-2020 |
| United States | 110 | Western Institutional Review Board - Connexus | 25-Jun-2020 |
| United States | 111 | SUNY Downstate | 24-Jun-2020 |
| United States | 112 | Western Institutional Review Board - Connexus | 10-Jul-2020 |
| United States | 113 | Western Institutional Review Board - Connexus | 10-Jul-2020 |
| United States | 114 | St. Joseph Health IRB | 02-Jul-2020 |
| United States | 115 | Providence St. Joseph Health | 02-Jul-2020 |
| United States | 116 | Providence St. Joseph Health | 02-Jul-2020 |
| United States | 117 | Western Institutional Review Board - Connexus | 07-Aug-2020 |
| United States | 118 | Western Institutional Review Board - Connexus | 13-Jul-2020 |
| United States | 120 | Western Institutional Review Board - Connexus | 29-Jun-2020 |
| United States | 123 | Providence St. Joseph Health | 25-Jun-2020 |
| United States | 124 | Western Institutional Review Board - Connexus | 22-Jul-2020 |
| United States | 125 | Western Institutional Review Board - Connexus | 15-Jul-2020 |
| United States | 126 | Western Institutional Review Board - Connexus | 27-Jul-2020 |
| United States | 127 | Western Institutional Review Board - Connexus | 25-Aug-2020 |
| United States | 128 | Western Institutional Review Board - Connexus | 04-Nov-2020 |
| United States | 129 | Western Institutional Review Board - Connexus | 15-Sep-2020 |
| United States | 130 | Western Institutional Review Board - Connexus | 30-Sep-2020 |
| United States | 132 | Western Institutional Review Board - Connexus | 18-Nov-2020 |
| United States | 133 | Western Institutional Review Board - Connexus | 20-Nov-2020 |
| Italy | 252 | ASST Grande Ospedale Metropolitano Niguarda Comitato Etico Milano Area C | 18-Aug-2020 |
| Spain | 275 | CEIm Instituto de Investigación Sanitaria La Fe | 19-Jun-2020 |
| Spain | 276 | CEIm Instituto de Investigación Sanitaria La Fe | 19-Jun-2020 |
| Spain | 277 | CEIm Instituto de Investigación Sanitaria La Fe | 19-Jun-2020 |
| Spain | 278 | CEIm Instituto de Investigación Sanitaria La Fe | 19-Jun-2020 |
| Spain | 279 | CEIm Instituto de Investigación Sanitaria La Fe | 28-Sep-2020 |
| United Kingdom | 300 | South Central - Oxford A | 30-Jun-2020 |
| United Kingdom | 301 | South Central - Oxford A | 30-Jun-2020 |
| United Kingdom | 302 | South Central - Oxford A | 07-Jul-2020 |
| Germany | 325 | Ethik-Kommission der Medizinischen Fakultät | 14-Jul-2020 |
| Germany | 326 | Ethik-Kommission der Medizinischen Fakultät | 14-Jul-2020 |
| Germany | 327 | Ethik-Kommission der Medizinischen Fakultät | 14-Jul-2020 |
| Germany | 328 | Ethik-Kommission der Medizinischen Fakultät | 14-Jul-2020 |
| Mexico | 350 | Hospital Universitario "Dr. Jose Eleuterio Gonzalez" | 03-Jun-2020 |
| Mexico | 352 | ITESM Campus Monterrey | 02-Jun-2020 |
| Mexico | 353 | Instituto Nacional de Cancerologia | 23-Jun-2020 |
| Mexico | 354 | Instituto Nacional de Cancerologia | 18-Jun-2020 |
| Mexico | 355 | Instituto Nacional de Cancerologia | 27-Jul-2020 |
| Mexico | 356 | Medica Sur | 28-Aug-2020 |
| Mexico | 357 | Instituto Nacional de Cancerologia | 18-Sep-2020 |
| Argentina | 375 | Stamboulian- Comite de Etica en Investigacion Clínica | 28-May-2020 |
| Argentina | 376 | Stamboulian- Comite de Etica en Investigacion Clínica | 09-Jun-2020 |
| Argentina | 377 | C.I.E.I.S. del Niño y del Adulto - Polo Hospitalario, Hospital Rawson | 17-Jun-2020 |
| Argentina | 378 | Comité Independiente de Ética de la Fundación Sanatorio Güemes | 22-Jun-2020 |
| Argentina | 380 | COMITÉ INSTITUCIONAL DE ÉTICA EN INVESTIGACIÓN H.Z.G.A.D. Evita Pueblo de Berazategui | 29-Jun-2020 |
| Argentina | 381 | Stamboulian- Comite de Etica en Investigacion Clínica | 24-Jun-2020 |
| Argentina | 382 | Comité de Investigación y Comité de Bioética del HIGA “Eva Perón” | 16-Jul-2020 |
| Argentina | 383 | Stamboulian- Comite de Etica en Investigacion Clínica | 02-Sep-2020 |
| Argentina | 384 | CEI de la Casa Hospital San Juan de Dios | 17-Sep-2020 |
| Argentina | 385 | Comité de Ética en Investigación Clínica y Maternidad Suizo | 11-Nov-2020 |
| Argentina | 386 | Stamboulian- Comite de Etica en Investigacion Clínica | 20-Oct-2020 |
| Brazil | 401 | Upeclin - Unidade de Pesquisa Clínica da Faculdade de Medicina de Botucatu - UNESP | 02-Jul-2020 |
| Brazil | 402 | Hospital Felício Rocho | 03-Jul-2020 |
| Brazil | 403 | Real e Benemerita Associação Portuguesa de Beneficiencia | 01-Jul-2020 |
| Brazil | 404 | CEPETI Centro de Ensino e Pesquisa em Terapia Intensiva | 10-Jul-2020 |
| Brazil | 405 | Hospital de Clinicas de Porto Alegre | 29-Jun-2020 |
| Brazil | 406 | Pesquisare | 06-Jul-2020 |
| Brazil | 407 | Hospital Santa Paula | 12-Jun-2020 |
| Brazil | 408 | Casa de Saude Santa Marcelina - Centro de Pesquisa Clinica | 30-Jun-2020 |
| Brazil | 409 | Hospital PUC-CAMPINAS | 29-Jun-2020 |
| Brazil | 410 | Hospital Alemão Oswaldo Cruz | 03-Jul-2020 |
| Brazil | 411 | COMITÊ DE ÉTICA EM PESQUISA DA LIGA NORTE RIOGRANDENSE CONTRA O CANCER | 07-Jul-2020 |
| Brazil | 412 | Faculdade de Medicina do ABC | 05-Aug-2020 |
| Brazil | 413 | CEMEC – Centro Multidisciplinar de Estudos Clinicos EPP Ltda | 30-Jul-2020 |
| Brazil | 415 | IPECC - Instituto de Pesquisa Clinica de Campinas | 31-Jul-2020 |
| Brazil | 416 | Hospital Carlos Fernando Malzoni Matao | 07-Oct-2020 |
| Brazil | 417 | Praxis Pesquisa Medica | 11-Oct-2020 |
| Brazil | 418 | CECIP - Centro de Estudos do Interior Paulista | 06-Oct-2020 |
| Brazil | 419 | Centro Hospitalar de Reabilitacao Ana Carolina Moura Xavier | 21-Sep-2020 |
| Russian Federation | 425 | First Moscow State Medical University n.a. Sechenov | 10-Jun-2020 |
| Russian Federation | 426 | Independent Moscow City Commitee | 17-Jun-2020 |
| Russian Federation | 429 | Saint-Petersburg City Pokrovskaya Hospital | 15-Jun-2020 |
| Japan | 450 | Tokyo Medical University Hachioji Medical Center | 26-Jun-2020 |
| Japan | 451 | Yokohama Municipal Citizen's Hospital | 03-Jul-2020 |
| Japan | 453 | Shintokai Yokohama Minoru Clinic IRB | 10-Jul-2020 |
| Korea, South | 475 | Ajou University Hospital | 17-Sep-2020 |
| Korea, South | 476 | Seoul Medical Center | 07-Oct-2020 |
| Korea, South | 477 | Seoul National University Boramae Medical Center | 21-Sep-2020 |
| Korea, South | 478 | Korea University Ansan Hospital | 22-Sep-2020 |
| India | 502 | Aakash Healthcare Super Speciality Hospital | 06-Nov-2020 |
| India | 503 | Government Medical College | 11-Dec-2020 |
| India | 504 | Government Medical College (GMC) Aurangabad | 11-Nov-2020 |
| India | 505 | Ruby Hall Clinic and Grant Medical Foundation | 04-Nov-2020 |
